# Effectiveness and Safety of Chloroquine or Hydroxychloroquine as a mono-therapy or in combination with Azithromycin in the treatment of COVID-19 patients: Systematic Review and Meta-Analysis

**DOI:** 10.1101/2020.07.25.20162073

**Authors:** Ramy Mohamed Ghazy, Abdallah Elmaghraby, Ramy Shaaban, Ahmed Kamal, Hatem Beshir, Amr Moursi, Ahmed Ramadan, Sarah Hamed N. Taha

## Abstract

Many recent studies have investigated the role of either Chloroquine (CQ) alone, Hydroxychloroquine (HCQ) alone, or CQ/HCQ in combination with azithromycin (AZM) in management of the emerging coronavirus. This systematic review and meta-analysis of either published or preprint observational or interventional studies were conducted to assess the cure rate, duration of hospital stay, radiological progression, clinical worsening, need for mechanical ventilation, the occurrence of side effects, and mortality. A search of the online database through June 2020 was performed and examined the reference lists of pertinent articles for in-vivo studies only. Pooled relative risks (RRs), standard mean, of 95 % confidence intervals (CIs) were calculated with the random-effects model.

**Results:** The duration of hospital stay was shorter in the standard care in comparison with HCQ group, the standard mean of hospital stay was 0.57, 95% CI, and 0.20-0.94. Overall virological cure, or more specifically at day 4, 10, and 14 among patients exposed to HCQ did not differ significantly from the standard care [(RR=0.92, 95% CI 0.78-1.15), (RR=1.11, 95% CI 0.74-1.65), (RR=1.21, 95%CI 0.70-2.01), and (RR=0.98, 95% CI, 0.76-1.27)] respectively. Radiological improvement or clinical worsening was not statistically different between HCQ and standard care [(RR=1.11, 95% CI 0.64-1.65) and (RR=1.28, 95% CI 0.33-4.99)]. The need for mechanical ventilation (MV) was not significant between the HCQ group and the standard care (RR= 1.5, 95%CI 0.78-2.89). Side effects were more reported in the HCQ group than the standard care (RR=3.14, 95% CI 1.58-6.24). Mortality among HCQ was not affected by receiving HCQ (RR=3.14, 95% CI 1.58-6.24), meta-regression analysis revealed that country is a strong predictor of mortality. The duration of hospital stay among the HCQ and AZM didn’t differ significantly from the standard care (standard mean= 0.77, 95% CI 0.46-1.08). Despite virological cure and need for MV did not differ significantly [(RR= 3.23, 95% CI 0.70-14.97) and (RR=1.27, 95%CI 0.7-2.13)] respectively. Mortality among the HCQ+AZM was more significantly higher than among the standard care (RR= 1.8, 95% CI 1.19-2.27).

**Conclusion:** Despite the scarcity of published data of good quality, the effectiveness and safety of either HCQ alone or in combination with AZM in treating the pandemic of COVID-19 can’t be assured. Future randomized control trials need to be carried out to verify this conclusion.

**Registration:** PROSPERO registration number: CRD42020192084

## BACKGROUND

Coronavirus disease-2019 (COVID-19) is a serious health problem caused by the novel Coronavirus (nCOV) or Severe Acute Respiratory Syndrome Coronavirus 2 (SARS-CoV-2)^(1)^. SARS-COV-2 is a member of the Coronavirus family, a family which was previously responsible for Severe Acute Respiratory Syndrome (SARS) in 2002 and Middle East Respiratory Syndrome (MERS) in 2012.^(2)^

COVID-19 was emerged by the end of 2019 at Wuhan City in China and was notified by WHO to be a pandemic in March 2020. ^(3)^ Till the 19th of July, 2020, 14,444,995 COVID-19 cases and 605,225 deaths were reported worldwide. ^(4)^

Till now, there is no effective treatment for COVID-19. ^(5)^ Chloroquine (CQ) was initially reported to be effective against SARS-COV-2 and then Hydroxychloroquine (HCQ) followed.^(6)^ SARS-COV-2 is known to bind to human cells via the Angiotensin-Converting Enzyme 2 (ACE 2) receptor.^(7, 8)^ In-vitro studies showed that CQ and HCQ cause glycosylation of ACE2 receptor making cells to be refractory to SARS-COV-2 infection.^(8)^ This makes the drugs possible players in the treatment and even the prophylaxis against COVID-19.

Both drugs have also shown to have immunomodulatory effects.^(9)^ HCQ is now broadly used in autoimmune diseases such as Lupus and Rheumatoid Arthritis.^(9)^ This makes both drugs potentially effective in reducing the severity of COVID-19 through suppressing the immune system response to SARS-COV-2, which is now thought to be at least partly responsible for the severe forms of the disease.^(8)^

The safety of both drugs is also an important issue. Although both drugs are generally well-tolerated, high doses can be associated with severe side effects like myopathy, neuropathy, and cardiomyopathy.^(10)^ Retinopathy is a well-known side effect that is related to prolonged use.^(9)^ Usage of CQ or HCQ in critically ill patients can carry a higher risk of side effects, especially when combined with other drugs that carry a risk of QT interval prolongation increasing the risk of torsade’s de points. ^(11, 12)^

In-vivo studies showed contradictory results regarding CQ and HCQ in COVID-19. Firstly, Chinese researchers reported the efficacy of CQ against COVID-19. ^(13)^ Then, a French group reported the efficacy of HCQ added to Azithromycin (AZM) in decreasing viral load.^(14)^ After that, many studies were reported showing no benefit or even harmful effects.^(12)^ Here, we conducted an in-vivo meta-analysis of efficacy and safety of CQ and HCQ in COVID-19.

## METHODS

We performed this systematic review in strict compliance with the preferred reporting items of the systematic review and meta-analysis PRISMA checklist ^(15)^. All steps were conducted in concordance with the Cochrane Handbook of Systematic Review and Meta-Analysis ^(16)^.

### Inclusion and Exclusion Criteria

#### Inclusion Criteria

Studies satisfying the following criteria were included:

- Recruited patients with confirmed SARS-COV-2 virus confirmed by Polymerase Chain Reaction (PCR).
- Declared the effect of CQ or HCQ as anti-SARS-COV-2.
- Had a comparator group receiving either standard care, placebo, and other antiviral treatment, with or without a control group.
- Reported any of the following outcomes:
  ○ Clinical improvement: the resolution of cough or fever.
  ○ Virological cure (proportion of virological cure either overall or at certain time day 4,10, or 14, or the number of days till virological clearance).
  ○ Laboratory test improvement (serum ferritin, lymphocyte count).
  ○ Hospital stay or number of days’ till discharge.
  ○ Radiological improvement.
  ○ Progression of clinical symptoms.
  ○ Need for mechanical ventilation (MV).
  ○ Death.
  ○ Safety of CQ and HCQ; reporting side effects and QT prolongation.
  ○ Duration of hospital stay, need for MV, virological cure rate, and mortality of the combination (HCQ and AZM).
- No restriction regarding country, race, gender, or age.

#### Exclusion Criteria

Any study had one of the following criteria would be excluded:

- Published before 2019.
- Conducted in non-human subjects or in vitro studies.
- Abstract-only papers as preceding papers, conference, editorial, and author response and books.
- Studies with data not reliably extracted, duplicate, or overlapping data.
- Any study was written in any language other than English, French, or Chinese.
- Case reports, case series, and systematic review studies were also excluded.

#### Comparisons

- HCQ or CQ in comparison to standard care.
- HCQ+AZM in comparison to standard care.

### Outcomes Conceptualization

#### Virological Cure Rate

The virological cure in this study includes the number of days until the PCR becomes negative. It includes also the virological cure rate in the days matched between at least two studies. Based on the results, we found matches on days 4, 10, and 14.

#### Hospital Stay

Hospital stay in this study is the duration of patients stays in hospital measured in days.

#### Radiological Progression

The radiological progression here includes the number of patients who show progression in their radiological CT results during the period of a study.

#### Clinical Worsening

By clinical worsening, we mean deterioration of the case during the study’s period, or development of complications such as severity progression, or worsening of clinical symptoms.

#### Need for Mechanical Ventilation (MV)

We mean by the need for MV the percentage of patients who needed respiratory support through MV during a study.

#### The Occurrence of Side Effects

This includes any side effect that happens from using the studied treatment during a study.

#### Mortality

Mortality in this study is the percentage of deaths that occur during a study period.

#### QT Prolongation

In this study, we target specifically the effect of HCQ/CQ/AZ on QT prolongation during a study period.

### Data Extraction

A computer literature searches of (PubMed, Google Scholar, Cochrane, Scopus, Web of Science, Segle, VLH, COVID-Inato, COVID-Trial-Clinical Trial.gov) was conducted till June 5^th^, 2020 using the following keywords (Chloroquine OR Hydroxychloroquine) AND (2019 novel Coronavirus disease OR COVID-19 OR SARS-CoV-2 OR novel Coronavirus infection OR 2019-ncov infection OR Coronavirus disease 2019 OR Coronavirus disease-19 OR 2019-ncov disease OR COV OR Coronavirus). (Eight independent authors screened the literature search results for relevant studies according to the pre-specified inclusion and exclusion criteria.

All records were collected into an Endnote library to find and delete the duplicates using the “remove duplicating function” with two options is mandatory. All references that had (1) same title and author and published in the same year, and (2) same title and author, and published in the same journal, would be deleted. References remaining after this step were exported to a Microsoft Excel file with essential information for screening. These include the authors’ names, publication year, journal, DOI, URL link, and abstract.

The title and abstract screening were done by seven independent reviewers to select papers based on inclusion criteria. Each article was checked by two independent reviewers. Any disagreement was solved by the first author (RG). During the full-text screening phase, all selected articles were downloaded, and the full text was reviewed by two independent reviewers. The decision to include or exclude articles for qualitative and quantitative analysis should be agreed by the two reviewers to pass through. If any disagreement was noticed, the first author was asked to give his decision. The completed data were then thoroughly checked by two reviewers (RMG, AK)

We applied three methods to do manual searching. Firstly, we searched the reference lists of the included articles. Secondly, we performed what is known as citation tracking in which the reviewers track all the articles that cite each one of the included articles. This might involve the electronic searching of databases. Thirdly, similar to the citation tracking, we followed all “related to” or “similar “articles. All excluded records were given exclusion reasons. Manually added research included preprint, and unpublished data if fulfilling the inclusion criteria.

During the data extraction and the quality assessment, in a Microsoft Excel sheet, two reviewers extracted data related to patient characteristics and outcomes (authors, year of publication, country of patients, inclusion or exclusion criteria, when the study was conducted, study’s design, sample size, treatment option, dosage and duration, adverse events, primary and secondary outcome). All collected articles and data extracted can be found here (Supplementary Data 1).

### Data Analysis

#### Method of Data Analysis

Data were analyzed using Review Manager Software V5.3 for Windows. For the continuous variables, data were pooled using the mean difference. For the categorical variables, data were pooled using Risk Ratio (RR) with the perspective of 95% Confidence Interval (CI) in the meta-analysis model. In the case of zero frequency, the correction value of 0.1 was used. In the case of significant heterogeneity, we used the random effect model, otherwise, the fixed-effect model was used. Meta-regression analysis was done to examine the impact of age difference on HCQ regimen group mortality RR.

#### Heterogeneity

Heterogeneity was assessed by the Chi-Square test (X^2^) and measured by the I-Square test. I-Square (I^2^) statistic was used for heterogeneity evaluation. Following Cochrane Handbook for Systematic Reviews of Interventions 10, I^2^ was interpreted as follows: “0% to 40%: might not be important; 30% to 60%: may represent moderate heterogeneity; 50% to 90%: may represent substantial heterogeneity; 75% to 100%: considerable heterogeneity. The importance of the observed value of I^2^ depends on (i) magnitude and direction of effects, and (ii) strength of evidence for heterogeneity (e.g. P-value from the chi-squared test, or a confidence interval for I^2^). In the case of heterogeneity, DerSimonian and Laird random-effects models were applied to pool the outcomes. Otherwise, the inverse variance fixed-effect model was used. In the case of missing standard deviation (SD), we calculated it from the corresponding 95% confidence interval or the standard error ^(17)^. Forest plots were presented to visualize the degree of variation between studies. In the case of the absence of mean and standard deviation, authors were emailed and asked for the required data, or they were calculated according to the formula mentioned in this research ^(18)^.

#### Quality Assessment

Quality assessment (QA) of the research depended on the study design. The risk of bias in the individual studies included for meta-analysis was assessed using the Cochrane risk assessment tool in cases of Randomized Control Trials (RCT) ^(19)^, study quality assessment tools for observational study ^(20)^, and Robins-1 for Non-Randomized control trial ^(21)^. The assessment was performed by two independent reviewers (AA, AK, SH) and further checked by two additional reviewers (RG, RS). The risk of bias for non-randomized control trial and observational study is found in the following link (Supplementary Data 2).

#### Sensitivity Analysis

Sensitivity analysis is known to be an essential part in systematic reviews with meta-analyses to determine the robustness of the obtained outcomes to the assumptions made in the data analysis ^(23)^. We conducted leave ne sensitivity analysis to examine the effect of studies that greatly influence the result, especially by their weight through excluding them from the meta-analysis.

## RESULTS

### Study Selection Process

A total of 4730 articles were found after searching for 12 different databases. Of this number, 1151 duplicates found by Endnote X8, and 472 were published before 2019. So, they were excluded. Title and abstract included for 3107 papers resulted in the exclusion of irrelevant papers (2394), retracted articles (15), and manually found duplicates (586). Eligibility screening included 112 articles. Finally, 23 papers were eligible, in addition to, 12 papers added manually of which (14) entered in meta-analysis. Figure 1 shows the PRISMA flowchart of the selection process.

**Figure 1.**
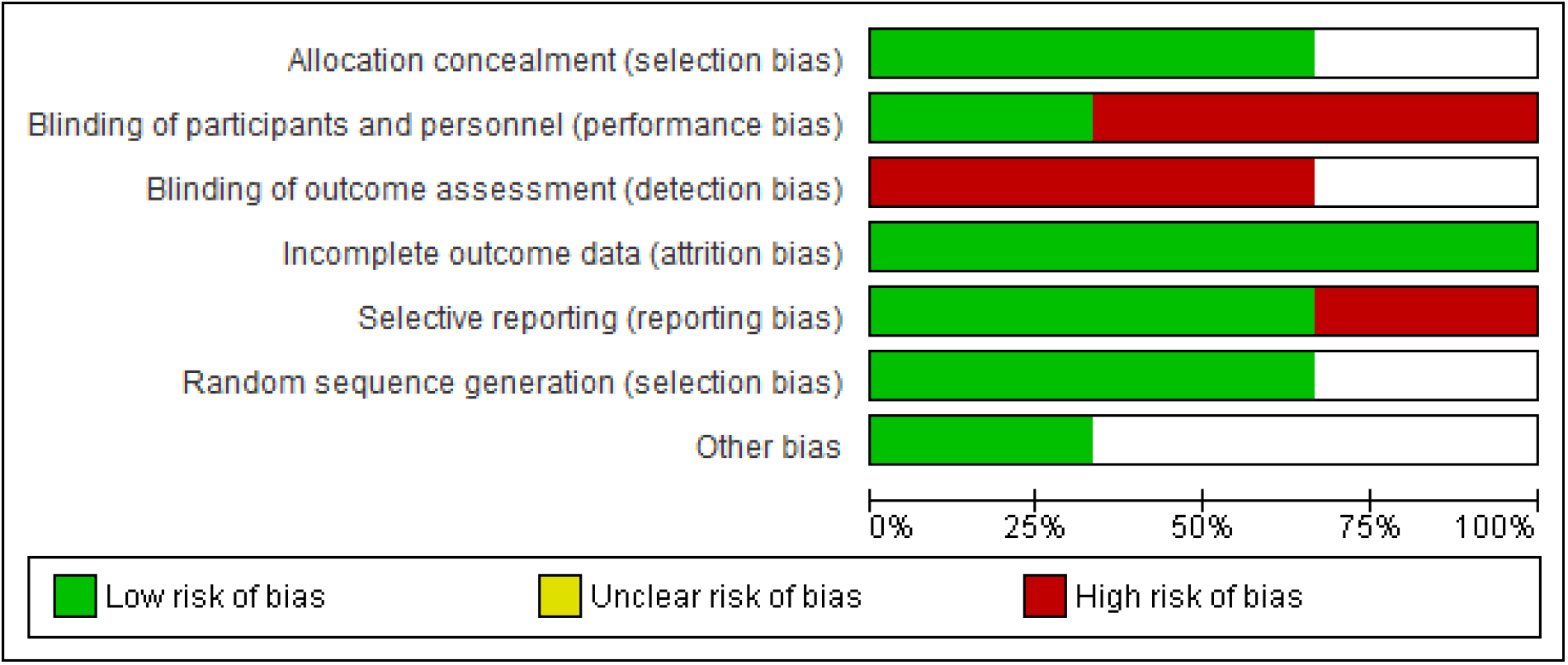
Publication Bias of Randomized control trial.

### Study Characteristics

Out of the fourteen studies entered into the meta-analysis, (2) studies were RCT (1) and non-RCT (3) case-control, and (8) retro or prospective cohort, of which, HCQ arms of the comparative studies have been combined with observational studies for effect size meta-analysis. The studies’ sample size ranged from (30 to 1438) participants. Characteristics of studies entered into the systematic review presented in Table 1.

**Table 1.**
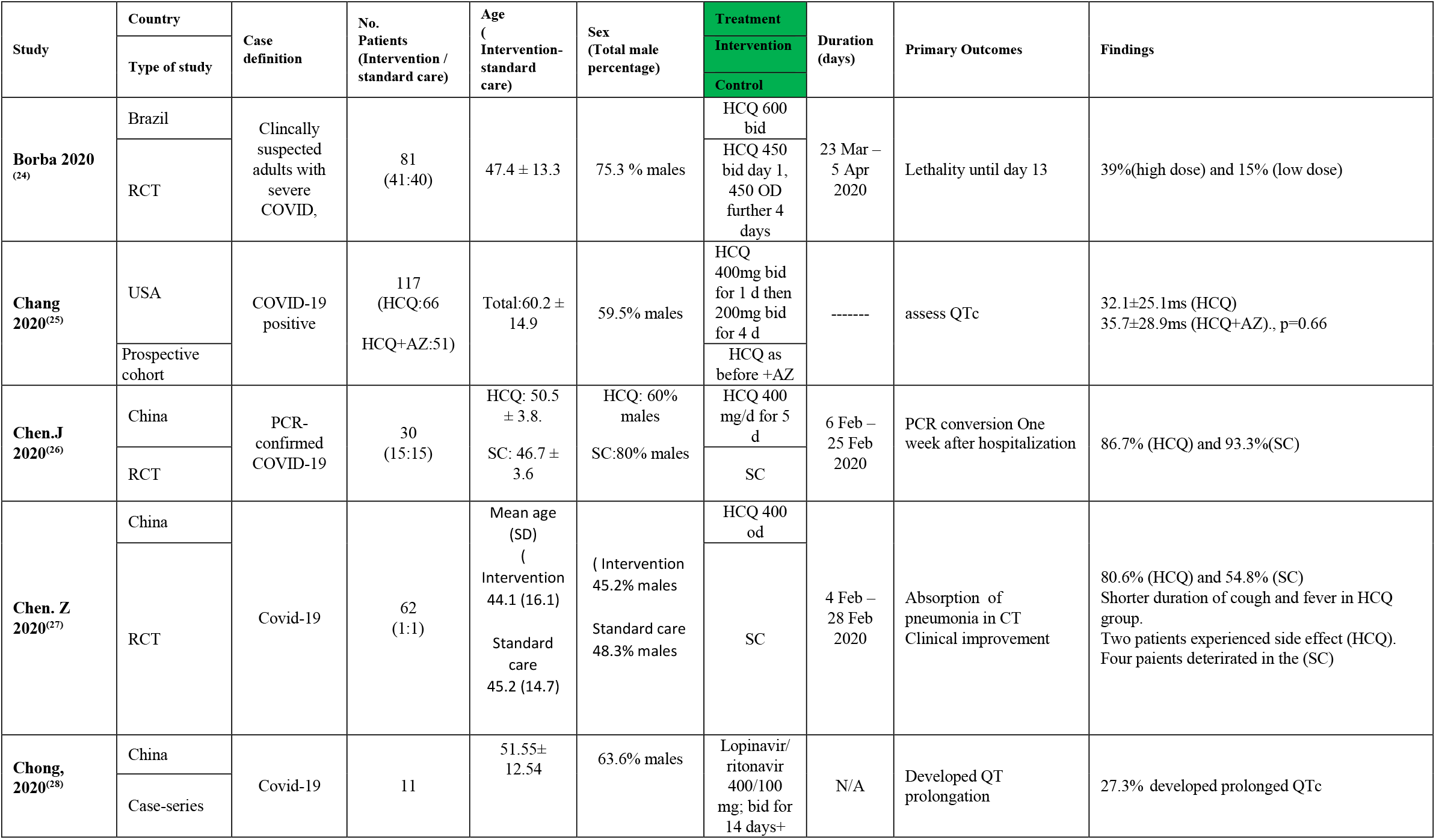
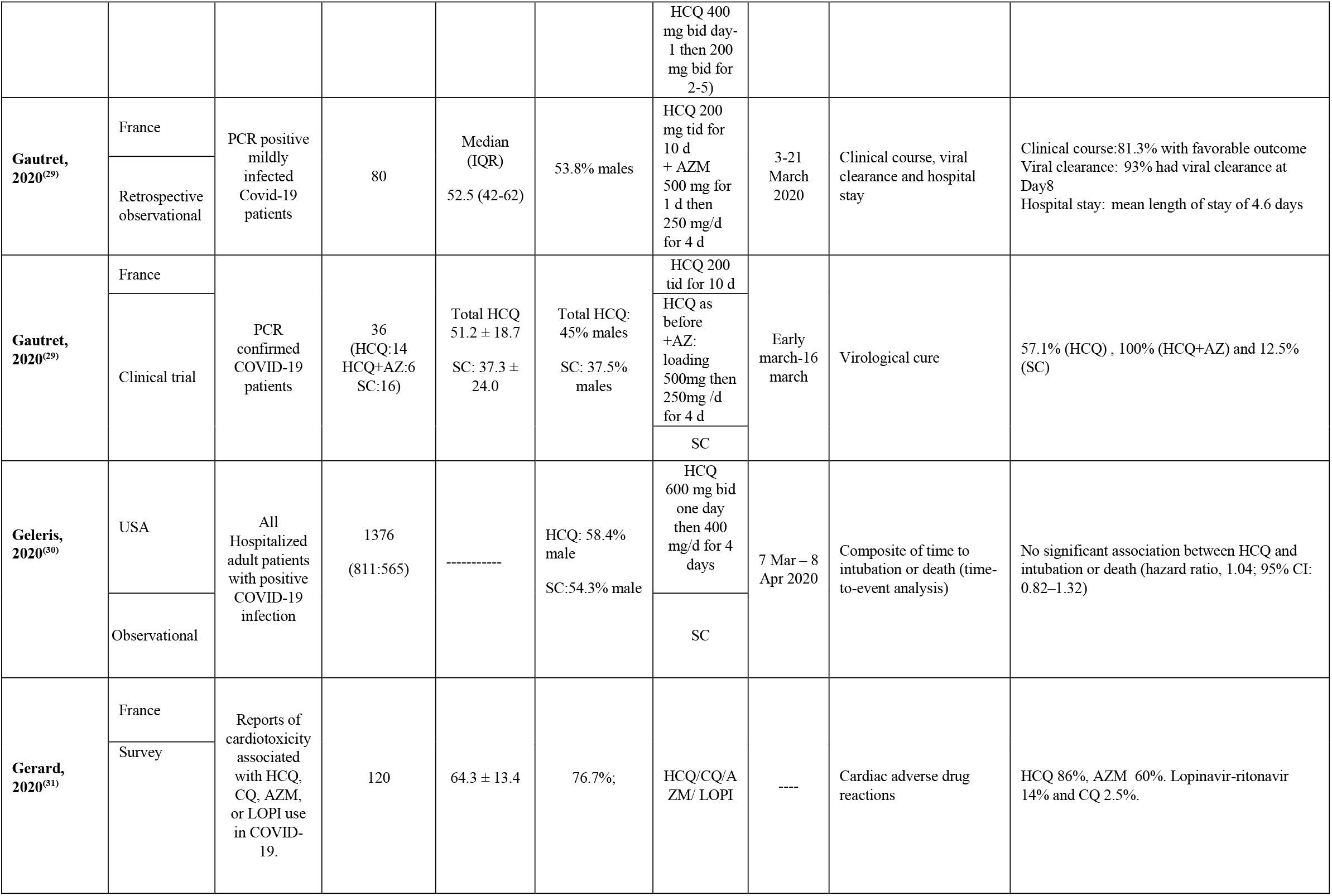
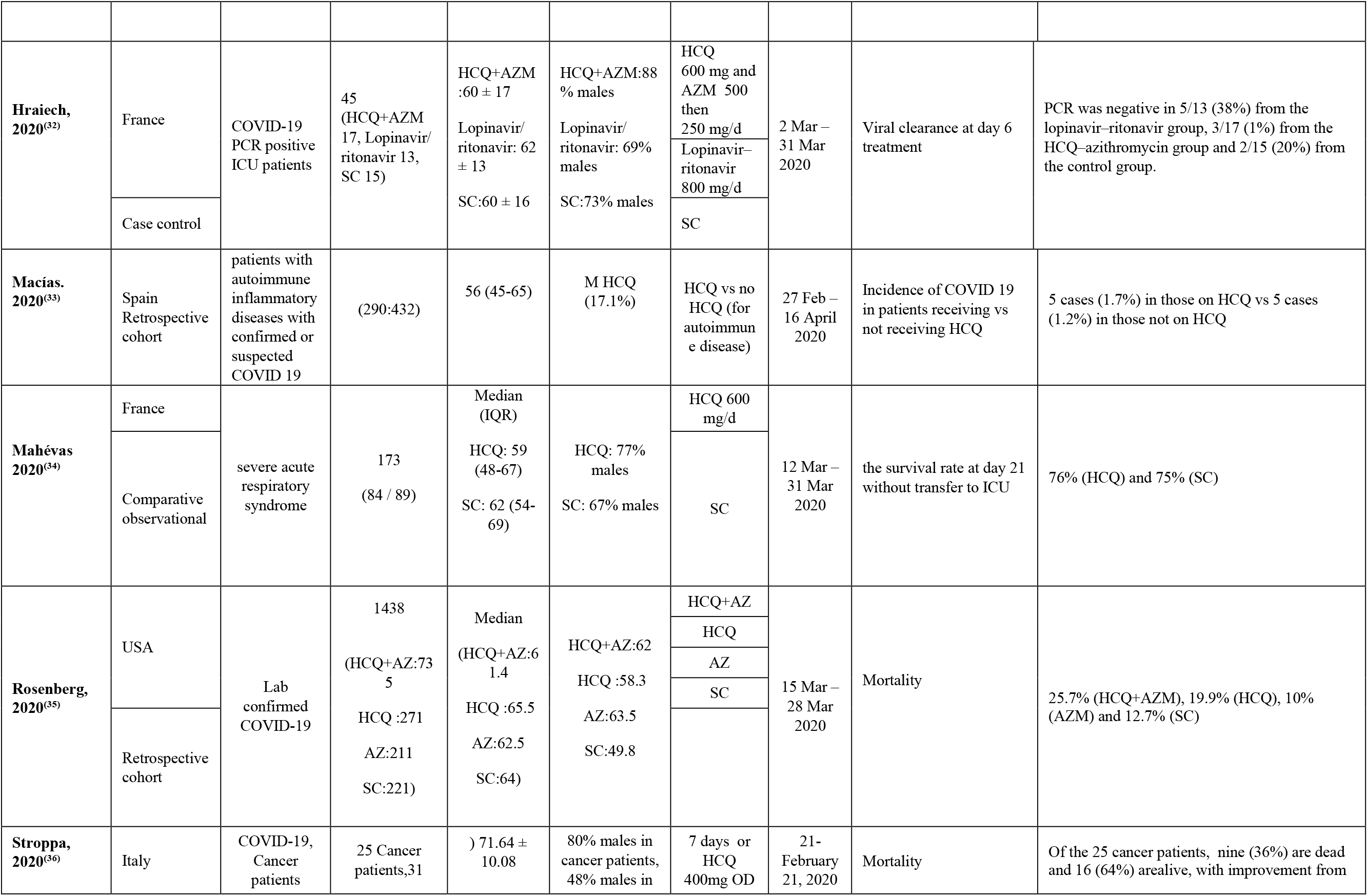
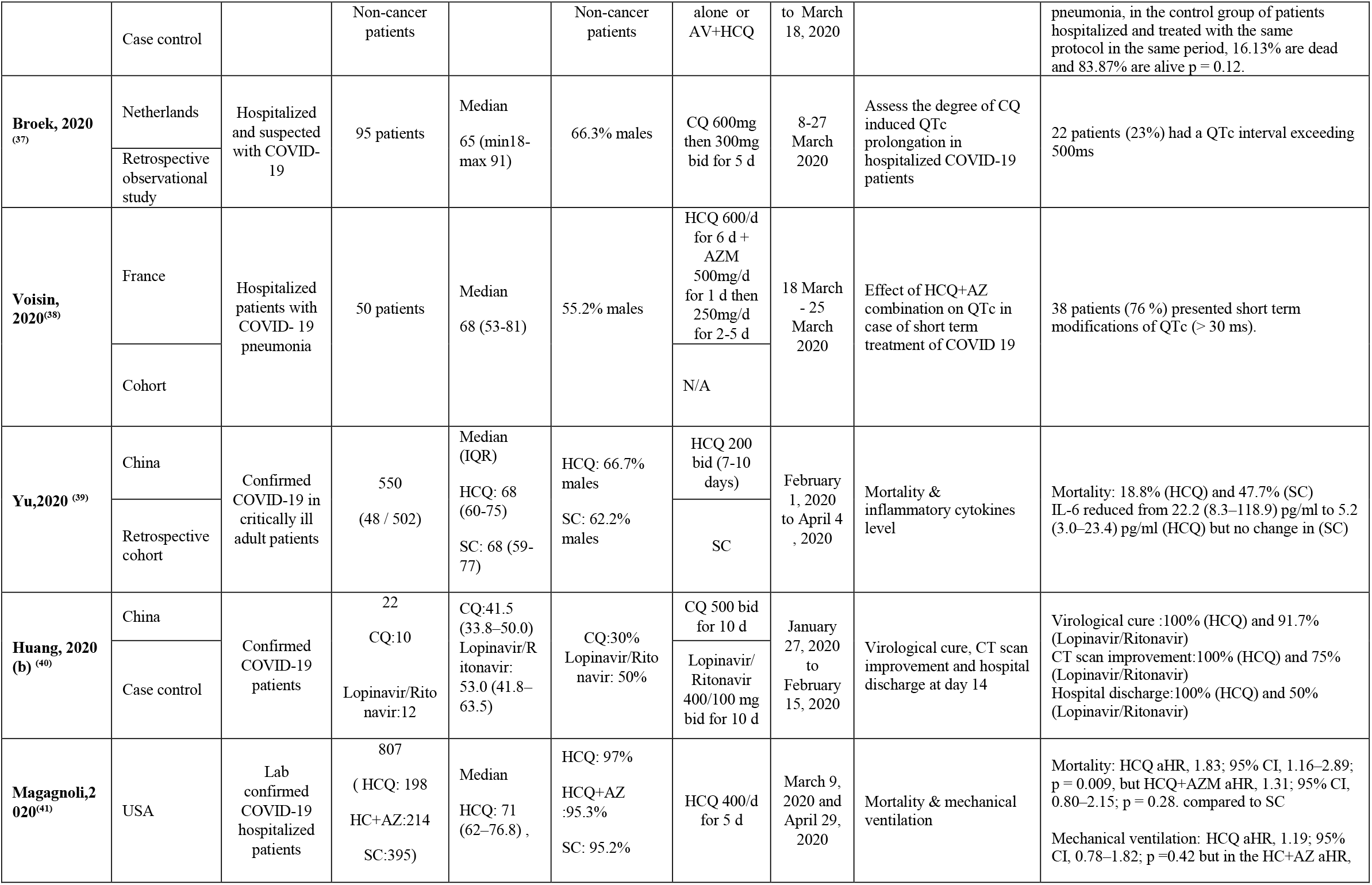
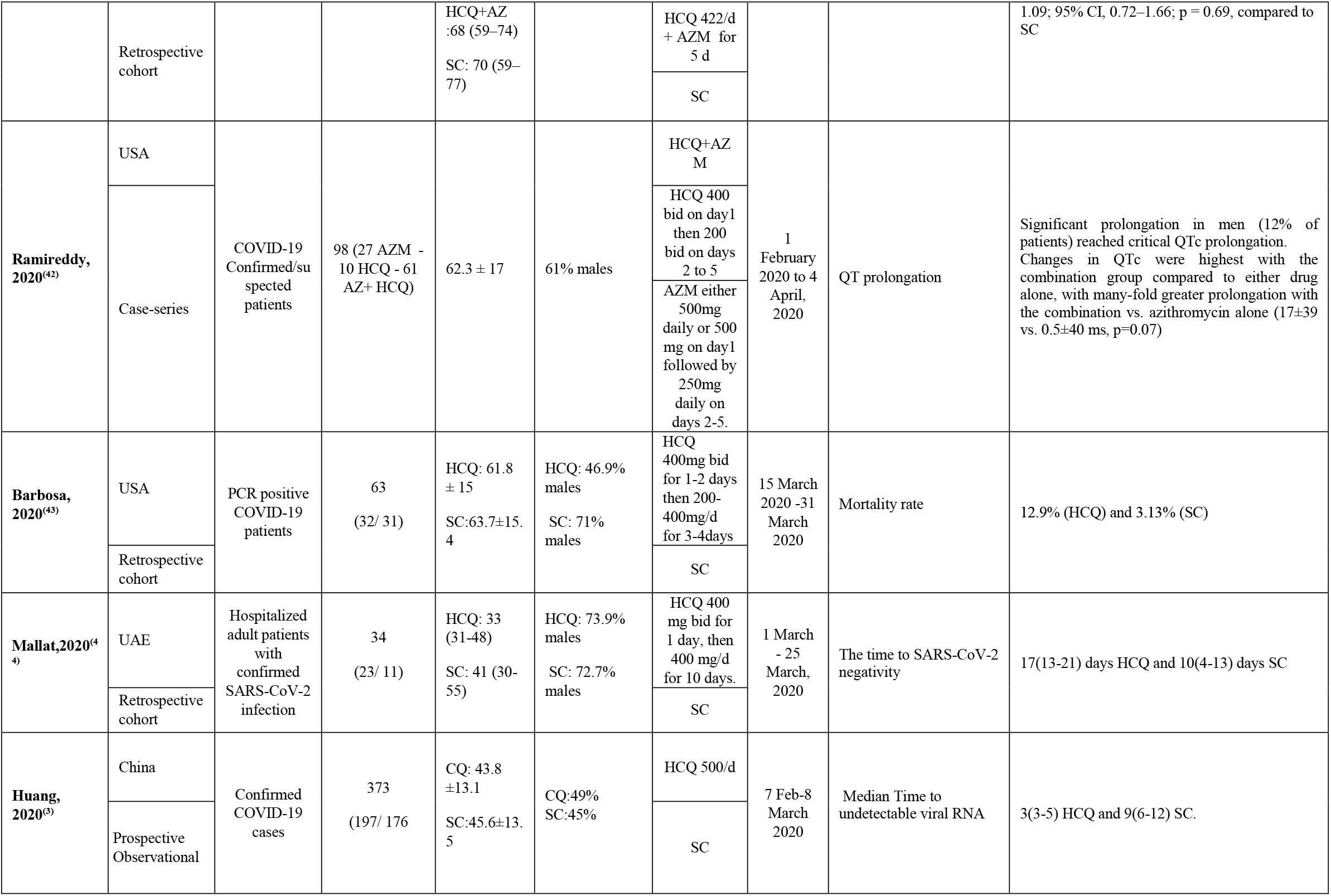
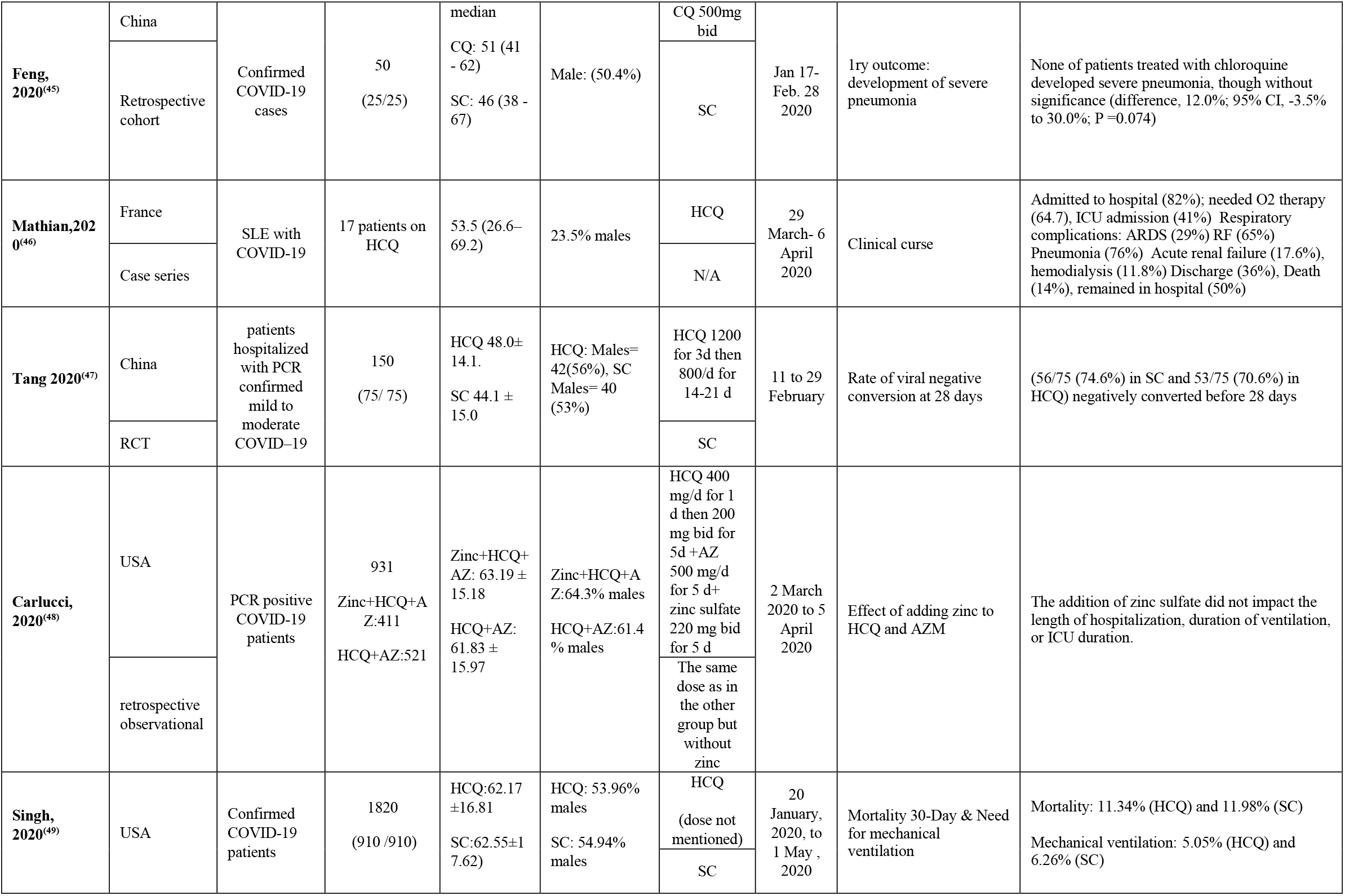
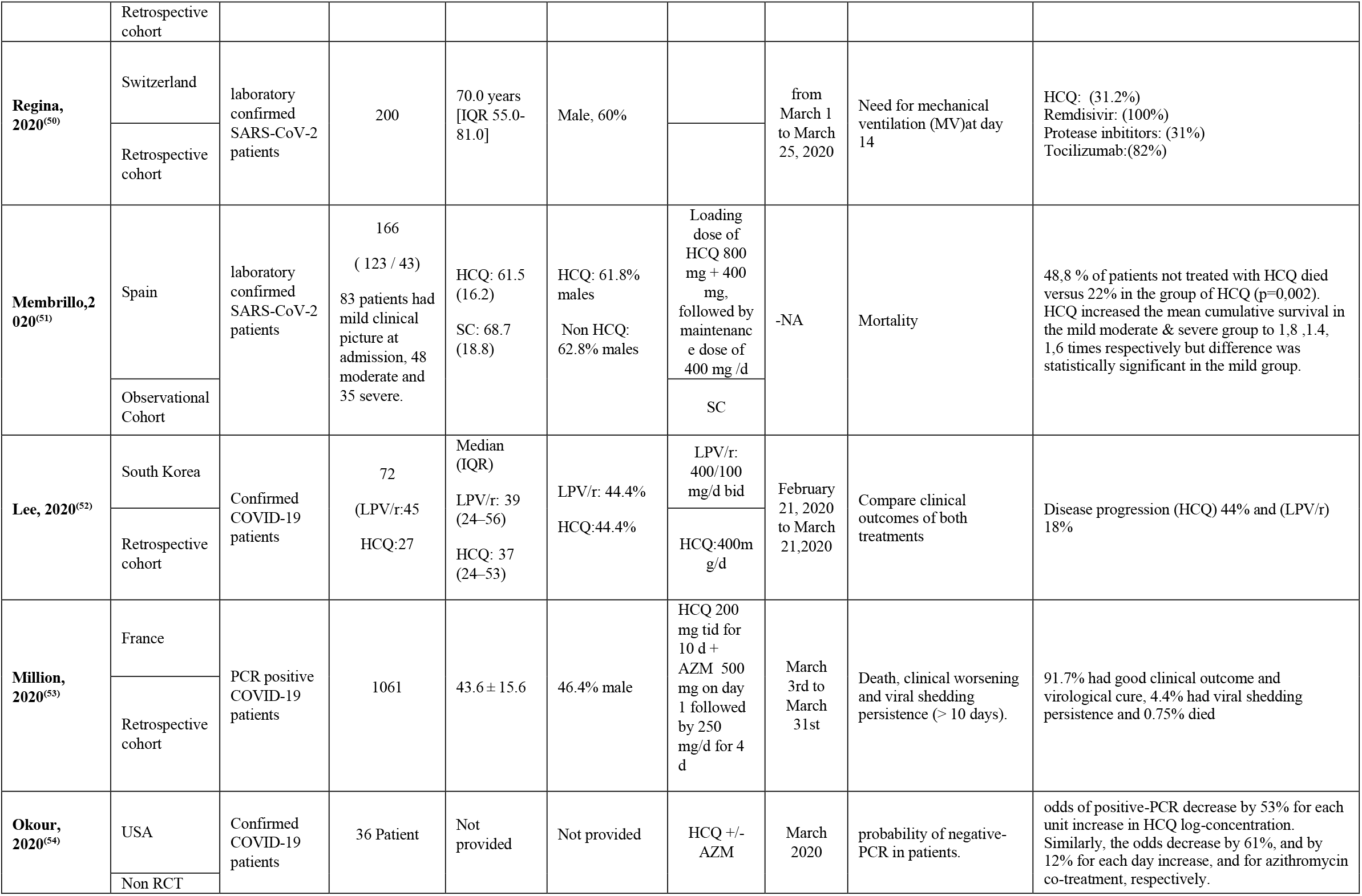
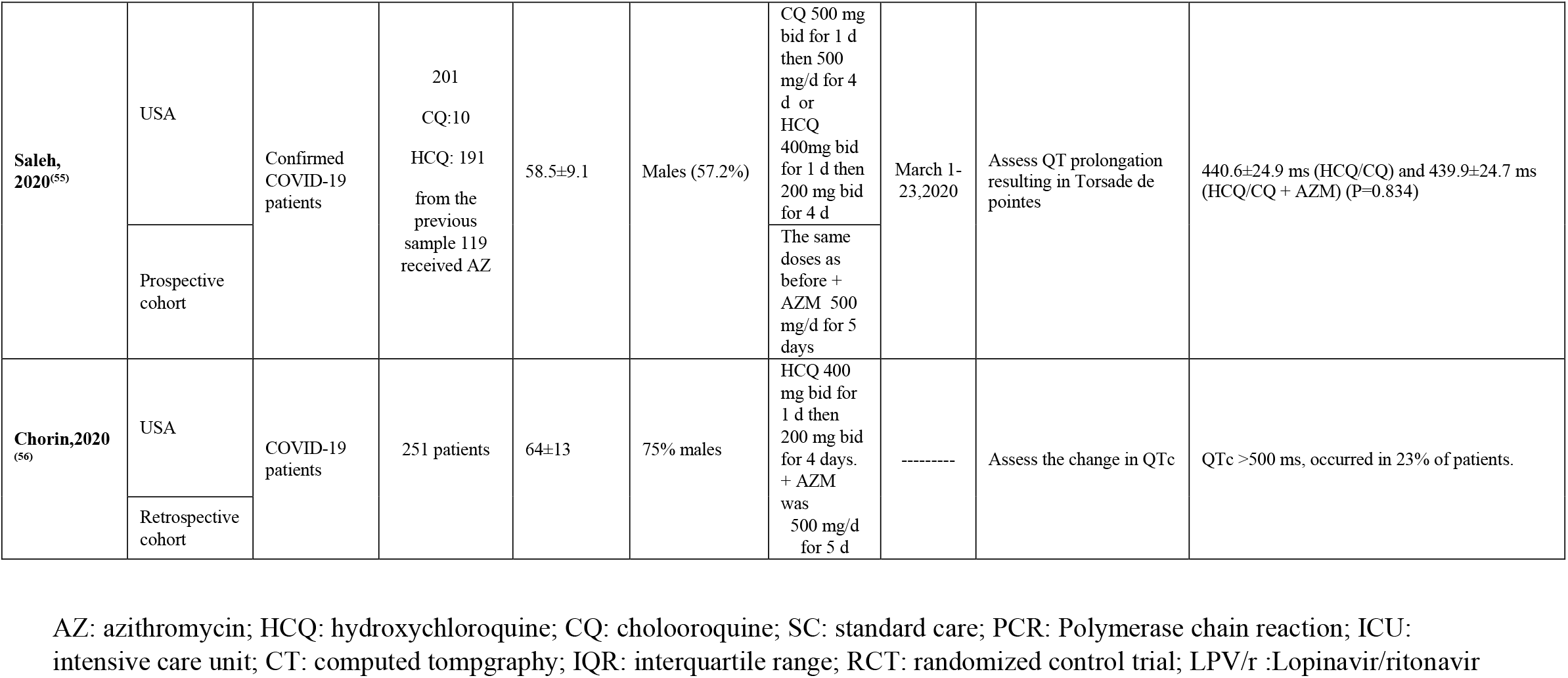
All published studies that reported the effectiveness or safety of Hydroxychloroquine, Chloroquine, or Azithromycin.

### Quality Assessment

Results of quality assessment for studies entered into a meta-analysis using Jadad, ROBINS-I, and NOS checklists were reported in the summary of the risk of bias has presented in Figure 2.

**Figure 2:**
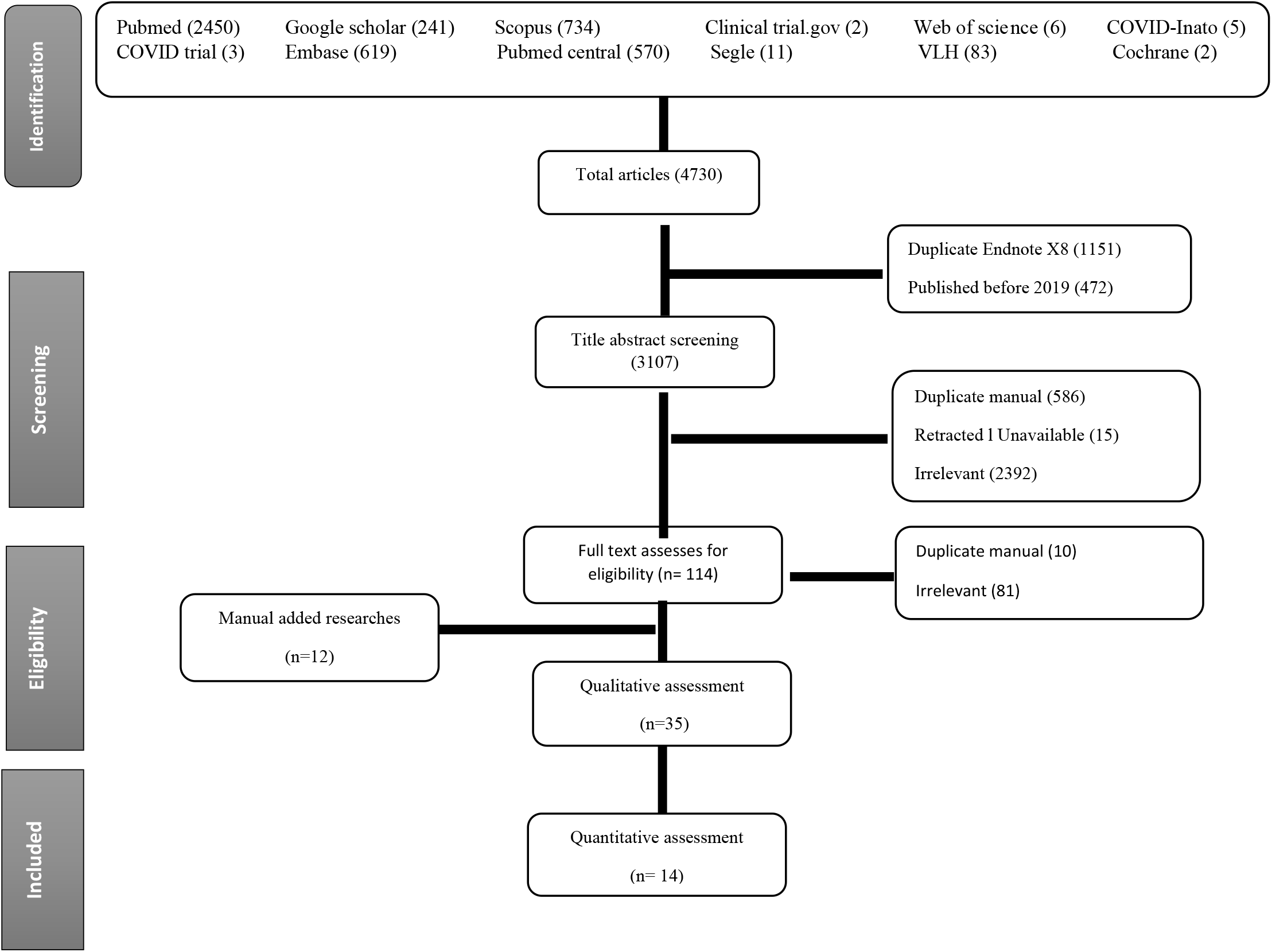
PRISMA flow chart of studies screened and included.

### Publication Bias

Publication bias assessment was conducted by visual inspection of the funnel plot. ^(22)^

### Hydroxychloroquine Versus Standard Care

#### Fever

A total of three studies that evaluated body temperature normalization after HCQ therapy; Huang et al,^(3)^ reported that body temperature returned normal after a geometric mean (coefficient variation), 1.2 (53.5) among HCQ group versus 1.9 (110.0) among non HCQ (P= 0.0029), while J.chen et al, ^(13)^ reported that patients’ temperatures returned to normal at approximately the same rate in both groups. (Median 1, IQR 0-2 for HCQ and Median 1, IQR 0-3 for no-HQR). Z. Chen et al,^(27)^ reported that the duration of fever was shorter in the HCQ group (mean 2.0±0.2) than in the non-HCQ group (mean 3.1±1.5).

#### Cough

Z. Chen et al, ^(27)^ reported that 15 of 31 (48.39%) of the control patients and 22 of 31 (70.97%) intervention patients had reported cough resolution. This difference was statistically significant P= 0.0016

#### Laboratory Test Improvement

Two studies evaluated the change in laboratory test after exposure to HCQ, first Mallat et al,^(44)^ reported that median lymphocyte count at day 7 was 1870 (1115-2625) compared to its baseline 1890 (1430-2230) in the control group, while it was 1650 (980-1950) at day 7 in the intervention group compared to its baseline level 1650(980-1950). Additionally, median serum ferritin level day 7 was 398 (52-1030) compared to its baseline 292 (33-1085) in the control group, while it was 249 (130-614) at day 7 in the intervention group compared to its baseline level 165 (63-320). Barbosa et al,^(43)^ reported that change in Neutrophil to Lymphocyte Ratio was higher in HCQ (9.55±21.5) versus standard care (1.58±6.26) but this increase was not significant. Similarly, the change in absolute lymphocyte count was not statistically significant between both groups (-0.61±0.52 of HCQ group vs −0.61±0.38 standard care).

#### Hospital Stay

The duration of the hospital stay of patients of the standard care group was significantly shorter than in the HCQ/CQ group (summary std. mean difference was 0.57, CI, 0.20-0.94). Of the four included studies, three studies favored the standard care with std. mean difference ranging from (0.50-1.19). The heterogeneity of the included studies was as follows (I^2^=92%, p <0.01). (Figure 3). In the sensitivity analysis, M. Huang, 2020 contributed most to heterogeneity. Excluding this study made the overall effect relatively higher (Z=12.33, P<0.000) and the test of heterogeneity was not significant (P = 0.54, I^2^=0%) (Figure 4)

**Figure 3.**
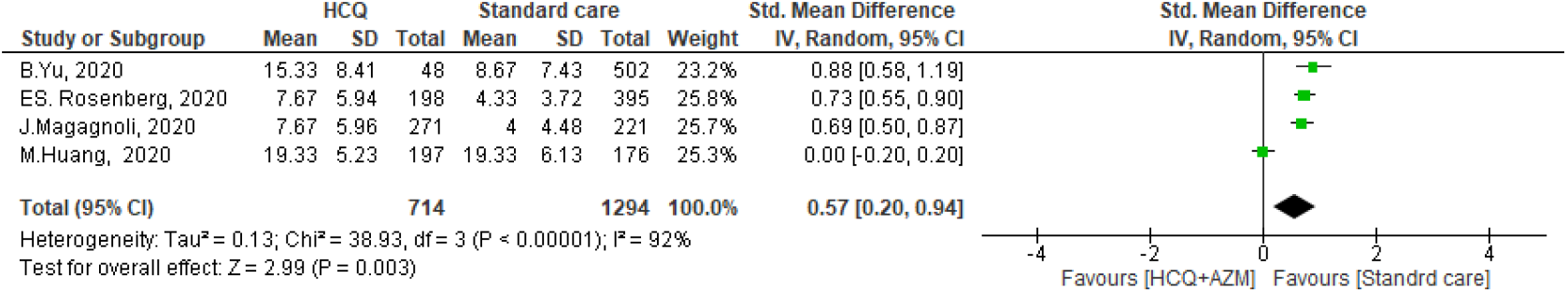
Duration of hospital stay of HCQ versus standard care.

**Figure 4.**
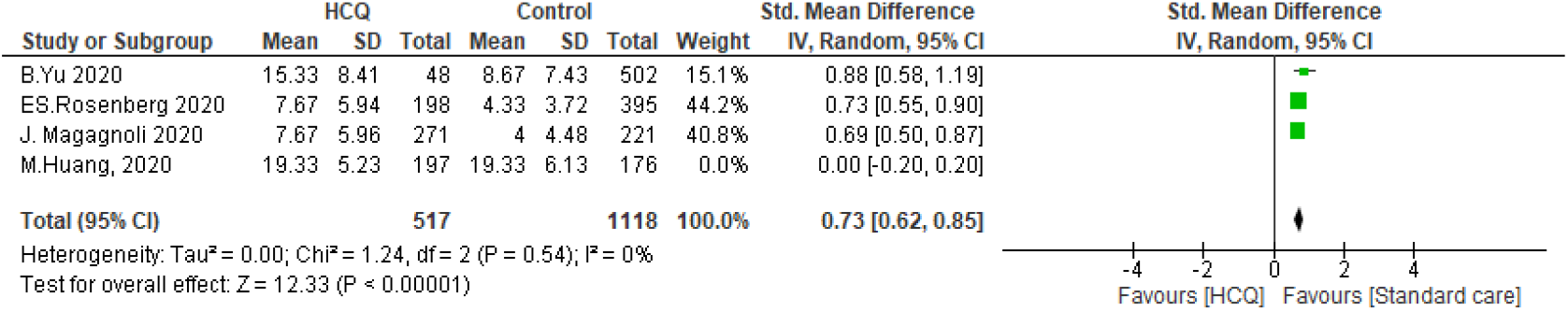
Sensitivity analysis by leave one out sensitivity analysis.

#### Virological Cure

##### Time to negative PCR

Four studies evaluated the time to negative PCR after administration of HCQ or CQ One study proved that intervention was more effective (Std. mean= -163, 95% CI -1.86 --1.39), however, the pooled Std. mean of these studies indicated there were no significant differences between the HCQ group and the standard care group in terms of the time for PCR to turn negative (RR: 0.05, 95% CI, -1.32-1.42). The measured heterogeneity was statistically significant I2=98%, P < 0.01. (Figure 5). Sensitivity analysis revealed that M. Huang, 2020 contributed most to heterogeneity. By the Exclusion of this study, the heterogeneity between the rest of the studies was insignificant (P=0.45, I^2^= 48%). Moreover, pooled std. mean turned to be significantly shorter in standard care groups (Z=2.32, P=0.02) (Figure 6).

**Figure 5.**
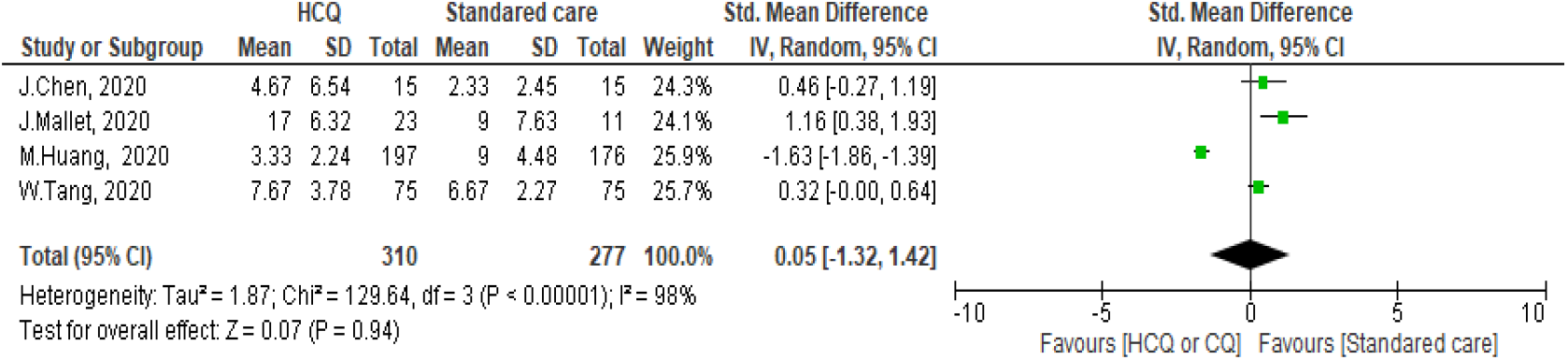
Forest plot for pooling risk ratios regarding the time to a negative PCR.

**Figure 6.**
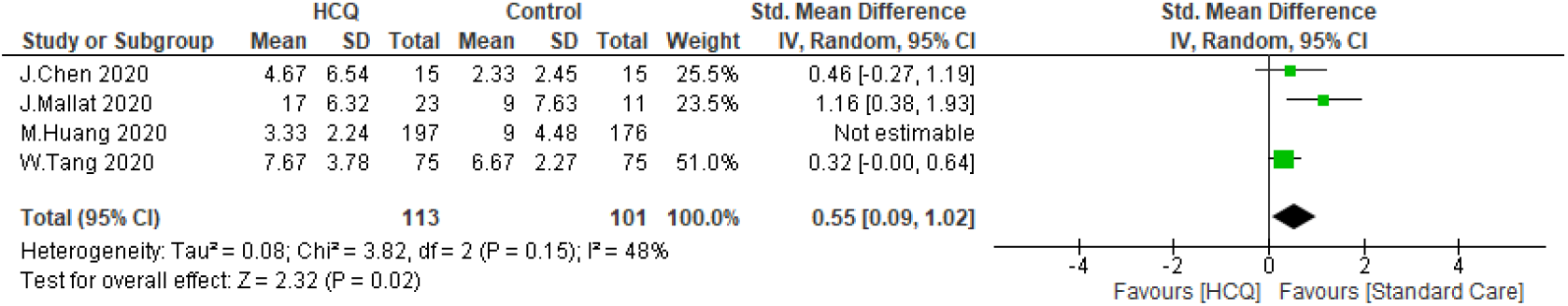
Sensitivity analysis by leave one out sensitivity analysis.

##### Virological cure rate

To get more insight over the virological cure, we were able to find two studies that analyzed the virological cure rate on day 4, two studies analyzed it on day 10, and three analyzed it on day 14. There were no differences between the HCQ group and the standard care group [(RR: 1.11, 95% CI, 0.26-4.69), (RR: 1.21, 95% CI, 0.70-2.10), and (RR: 0.98, 95% CI, 0.76-1.27)] (Figure 7-9). The heterogeneity of the three analysis were (I^2^ = 85%, P = 0.01, I^2^ = 95%, P < 0.01, and I^2^ = 85%, P < 0.01) respectively. The comparison of the virological cure rate at day 14 was subjected to leave one out sensitivity analysis. There was substantial heterogeneity between studies at all stages of the test. However, M. Huang, 2020 contributed to most of the heterogeneity (Figure 10).

**Figure 7.**
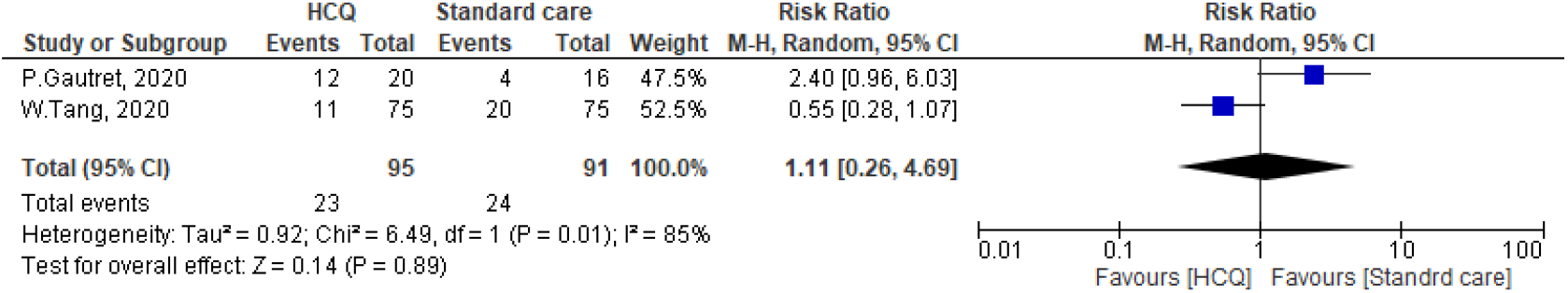
Forest plot for pooling risk ratios regarding virological cure rate on day four.

**Figure 8.**
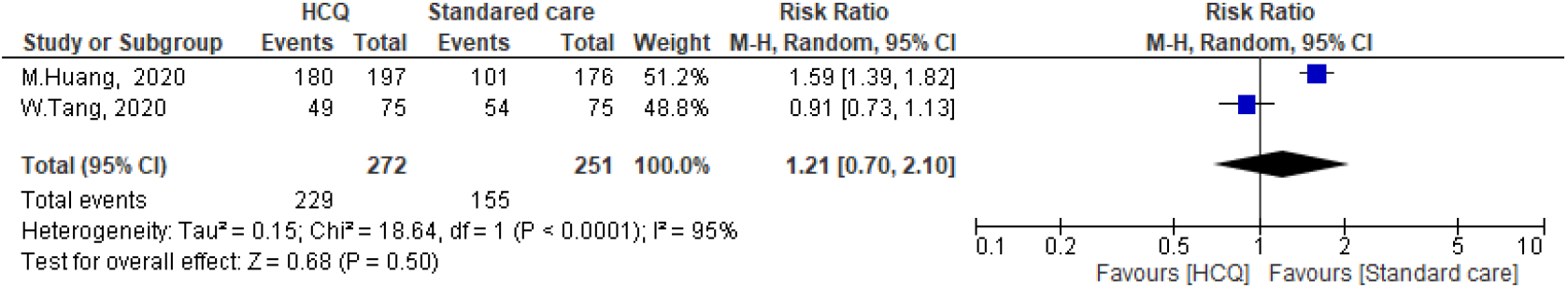
Forest plot for pooling risk ratios regarding virological cure rate on day 10.

**Figure 9.**
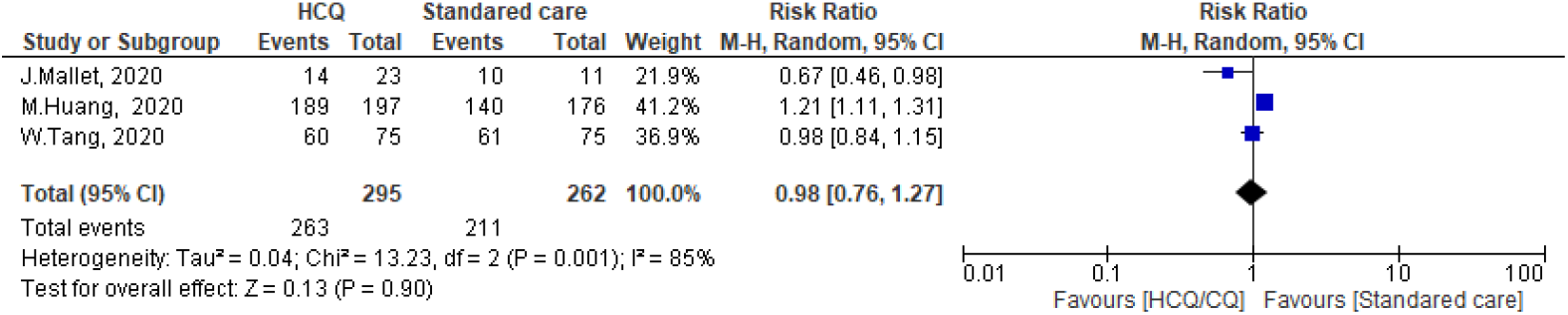
Forest plot for pooling risk ratios regarding virological cure rate on day 14.

**Figure 10.**
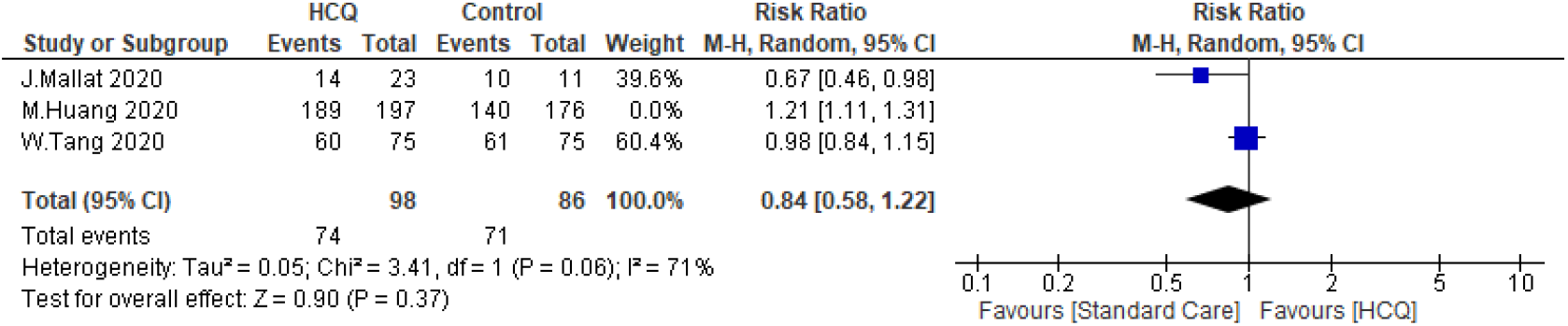
Leave one out sensitivity analysis.

The overall virological cure was not statistically significant between the intervention group and the standard care. The pooled RR was 0.91, 95% CI = 0.79-1.05). The heterogeneity of the studies was as follow I^2^ = 67%, P = 0.03 (Figure 12). In the sensitivity analysis, P. Gautret, 2020 contributed to most of the heterogeneity. By exclusion of this study, the heterogeneity was not significant between the rest of the studies (P=0.26, I^2^= 25%).

**Figure 11.**
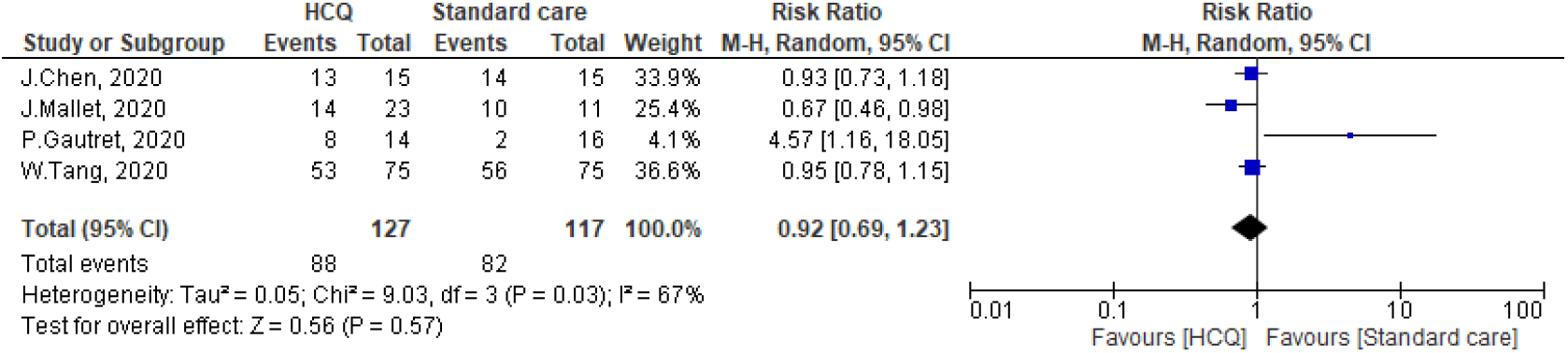
Forest plot for pooling risk ratios regarding the overall virological cure rate.

**Figure 12.**
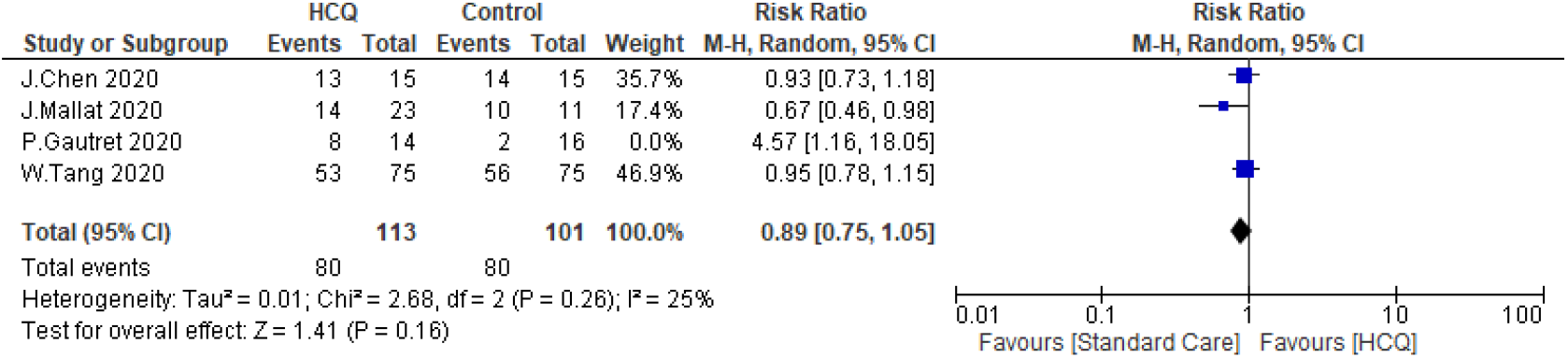
Sensitivity Analysis.

#### Radiological Improvement

Three studies evaluated the radiological improvement differences between the HCQ group and the standard care group. Only one study reported better radiological improvement of the standard care (OR=1.47: 95% CI, 1.02-2.11). The HCQ group didn’t statistically significantly differ from the standard care group (RR: 1.11, 95% CI, 0.74-1.65) (Figure 13). The heterogeneity between studies was not significant (P=0.16, I2=45%). Sensitivity analysis showed that Z Chen, 2020 contributes most of the heterogeneity. By exclusion of this study, the I^2^ index approached 0% (Figure 14).

**Figure 13.**
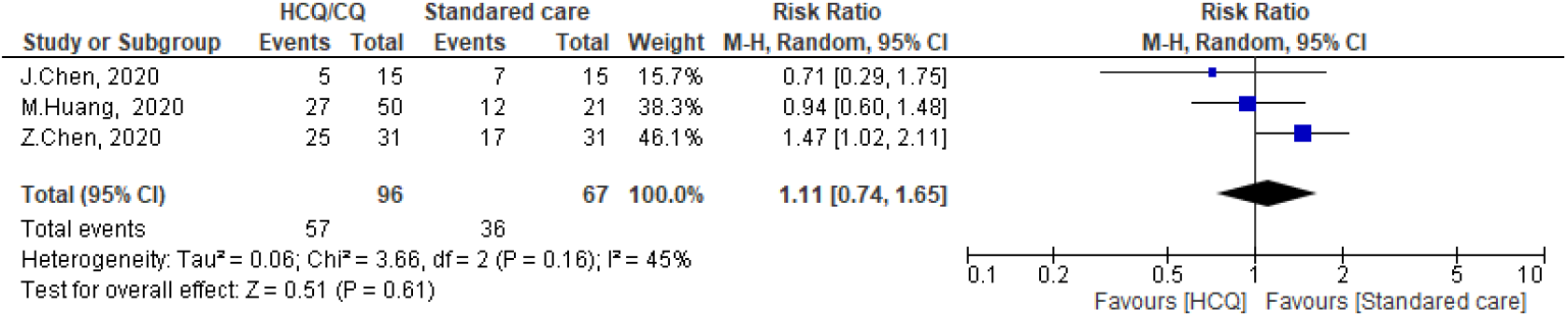
Forest plot for pooling risk ratios regarding radiological improvement.

**Figure 14.**
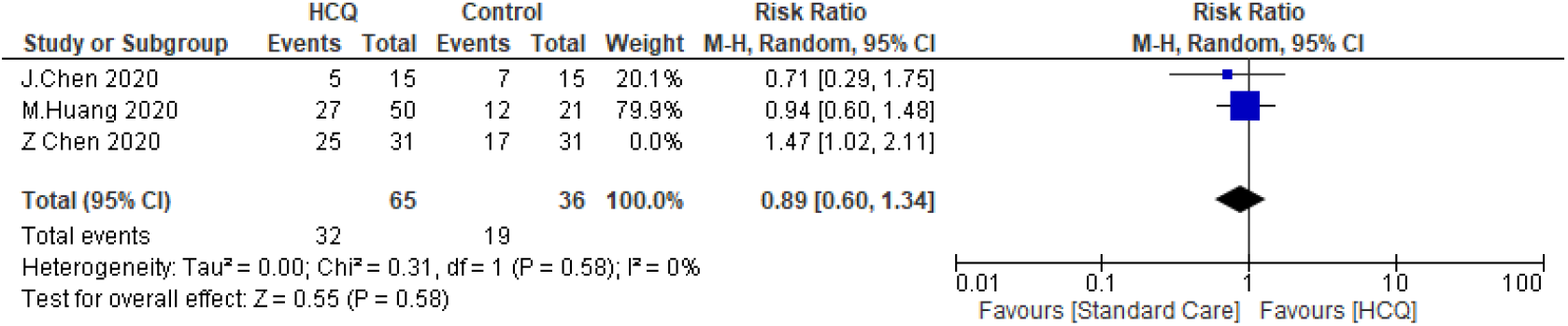
Leave one sensitivity analysis.

#### Worsening of Clinical Symptoms

Five studies evaluated the differences between the HCQ or CQ group and the standard care group in terms of clinical worsening. The meta-analysis showed there were no differences between the HCQ group and the standard care group regarding the worsening of clinical symptoms (RR: 1.28, 95% CI, 0.33-4.99). The heterogeneity of the studies was not significant P = 0.07 & I^2^ = 54% (Figure 15). Sensitivity analysis revealed that M. Huang, 2020 contributed most to heterogeneity. By exclusion of this study, the heterogeneity of studies was not significant (P = 0.29, I^2^ =19%) (Figure 16).

**Figure 15.**
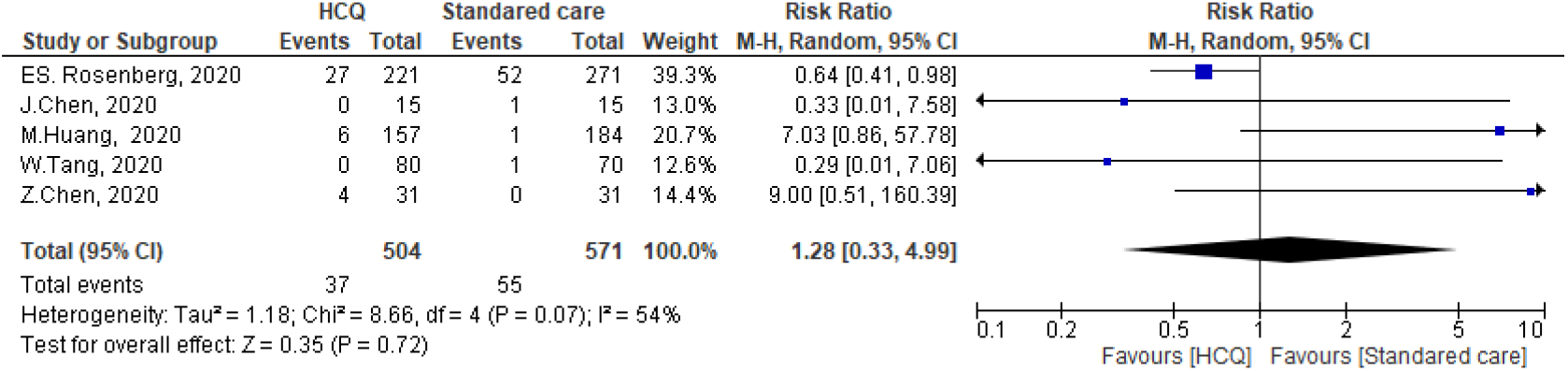
Clinical worsening of the hydroxychloroquine versus the standard care.

**Figure 16.**
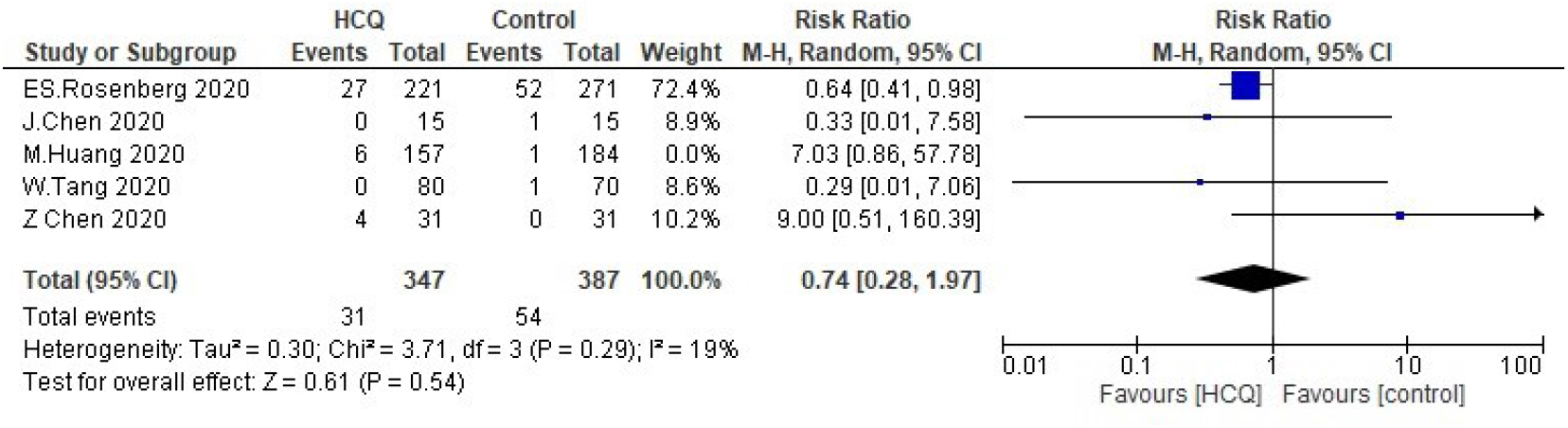
Leave one Sensitivity Analysis.

#### Need for Mechanical Ventilation

The need for MV was reported in five studies in both the HCQ group (118/1395) and the standard care group (156/1617). Two studies reported more need for mechanical ventilation among the standard care group, (rang of individual RR is 1.03-18.17, 95% CI). In the analysis, there was no significant difference between both groups (summary RR = 1.50, 95% CI, 0.78-2.89) as shown in (**Figure 17**). The test of heterogeneity was statistically significant I^2^=81%, P=0.001. Upon performing leave one out sensitivity analysis, there was a substantial heterogeneity at all stages of the test. However, ES. Rosenberg, 2020 contributed most to heterogeneity between studies. By exclusion of the study, the heterogeneity was insignificant (P=0.08, I^2^=61%) (Figure 18).

**Figure 17.**
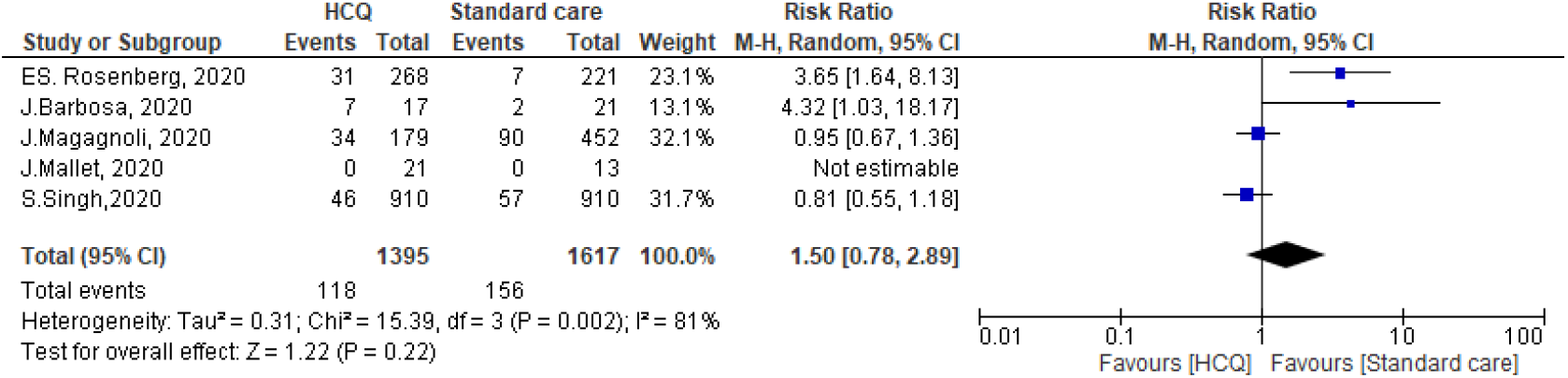
Need for mechanical ventilation of the hydroxychloroquine versus standard care.

**Figure 18.**
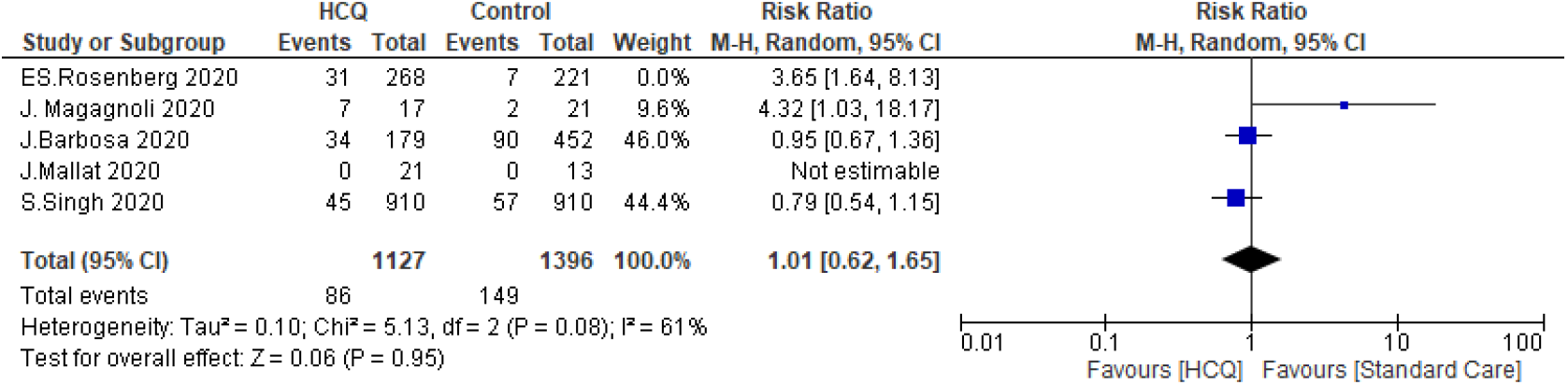
Leave one sensitivity analysis.

#### Side Effects

Three studies addressed side effect of HCQ revealed that intervention group witnessed greater side effects than the standard care (27/116) and (9/126) respectively, this difference was statistically significant (pooled RR= 3.14, 95% CI: 1.58-6.24). The heterogeneity of the studies was not significant (I2=0%, P=0.79) (Figure 19).

**Figure 19.**
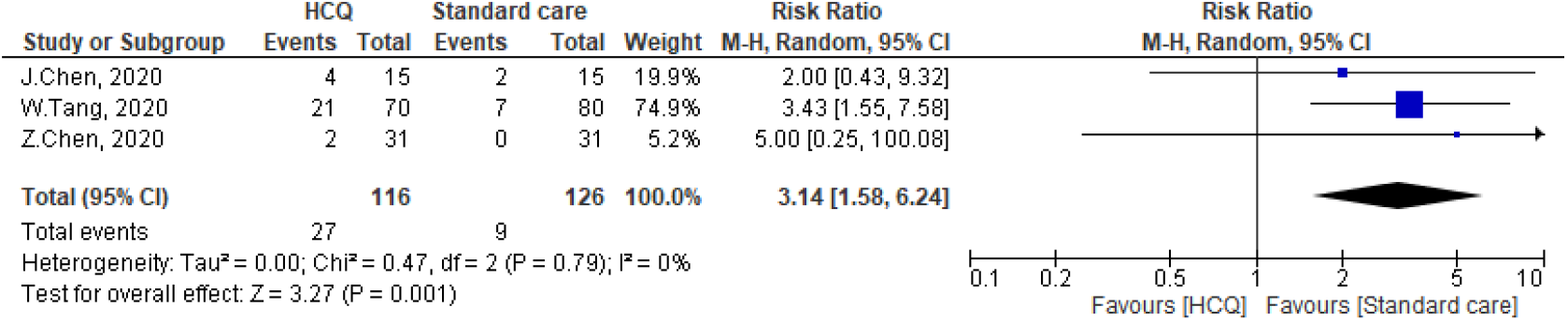
Side effects of the hydroxychloroquine versus the standard care.

#### QT Prolongation

Many studies have evaluated the effect of HCQ in inducing QT prolongation; Chong et al. demonstrated that 45.5% of patients exposed to HCQ developed QT prolongation. On the same vein, Broek et al, ^(37)^ noticed that 23% of CQ patients developed significant QT prolongation (>500msec). Voisin et al, ^(38)^ reported that of 50 patients treated with HCQ+AZM; 6 patients stopped the treatment due to significant QT prolongation, and 8 patients and 38 patients (76 %) presented short term modifications of QTc duration (meaning > 30 ms). The same figure was reported by Ramireddy et al. ^(42)^ and Saleh et al.,^(55)^ who reported that there is no difference regarding QT prolongation between patients treated with CQ or HCQ. Moreover, combination with AZM increased the risk of QT prolongation as (470.4±45.0 ms) versus monotherapy (453.3±37.0 ms), P=0.004. This increase in QT prolongation was incriminated in discontinuation of treatment in 3.5% of the studied patients. On the other hand, Rosenberg et al,^(35)^ reported a lower incidence of QT prolongation among patients treated with a combination of HCQ+AZM versus HCQ alone (11.0% vs 14.4%) respectively. Finally, Chang et al., ^(57)^reported that 17.9% of patients treated with HCQ± AZN had QT prolongation > 500 m-second. The prolongation of QT after administration of HCQ+AZM or HCQ alone was not significantly different.

#### Mortality

Mortality was addressed in 8 studies and controversial results were seen. B. Yu et al,^(39)^ FJ and Membrillo et al ^(51)^ showed that there is more mortality in the standard care groups in comparison to the HCQ. While, Rosenberg et al and Magagnoli et al,^(41)^ showed that there was more mortality in those who did receive HCQ. It is worthy to mention that, ‘among the 20 patients included in the study of Guatret et al, ^(14)^ 6 patients were on AZM In the analysis there was no significant difference between the two groups with RR of 0.99, 95% CI 0.61, 1.59 as shown in Figure 20. Leave one out sensitivity analysis revealed a considerable heterogeneity at all stages of the studies. All studies nearly equally contributed to the overall heterogeneity. Hence, meta-regression was needed to underline the possible effect of covariates. The risk of mortality was regressed considering mean or median age, country, percentage of male patients, and severity of illness as regressors. Age was not a significant predictor (P=0.323) as the mean or median age was above 60 years across all selected studies except for P. Gautret, et al., study ^(14)^ in which the median age of the participant was 45 years. Moreover, the severity of illness was not significant (P=0.105) as the patients in almost all selected studies were hospitalized with varied clinical status except for B. Yu, et al,^(39)^ in which the patients were all critically ill. Interestingly, the country of the study was a significant predictor for the risk of mortality at two levels (France and USA) setting for China (B. Yu, 2020) as a reference country. Switching from Chinese to French studies increases the relative risk of mortality in HCQ groups by 7.28 times (P=0.001) concerning SC groups. Similarly, switching from Chinese to American studies increased the relative risk of mortality in HCQ groups by 4.29 times (P=0.005). By Meta-regression, the overall heterogeneity of selected studies was not significant (0.243, I^2^=22%). Publication bias of included study is presented in (Figure 21).

**Figure 20:**
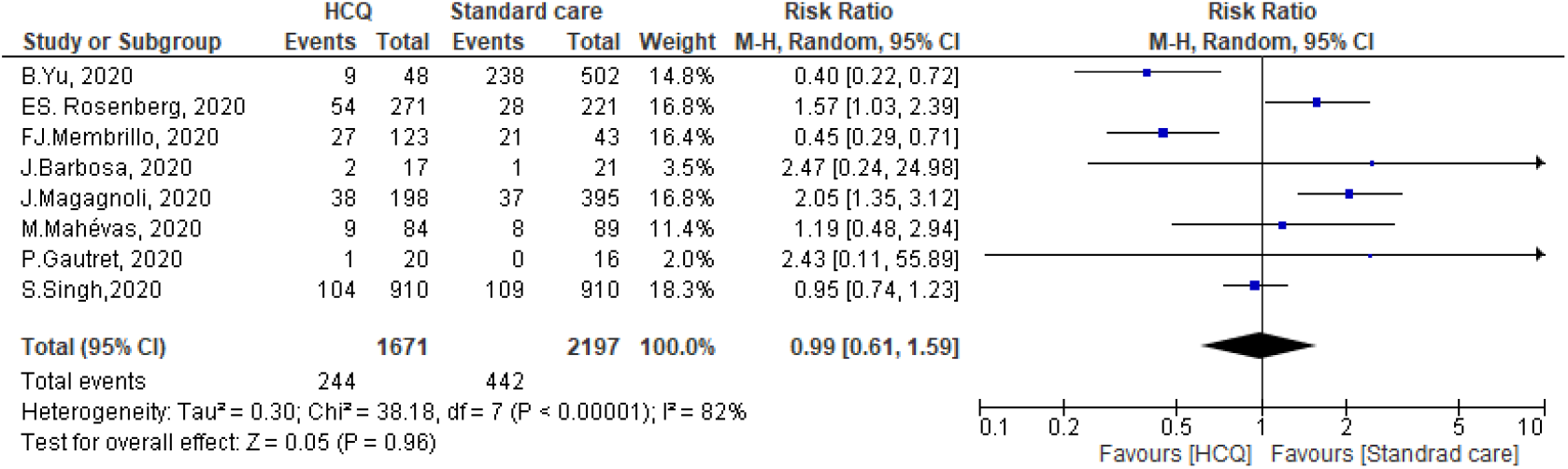
Mortality in the hydroxychloroquine vs standard care group.

**Figure 21.**
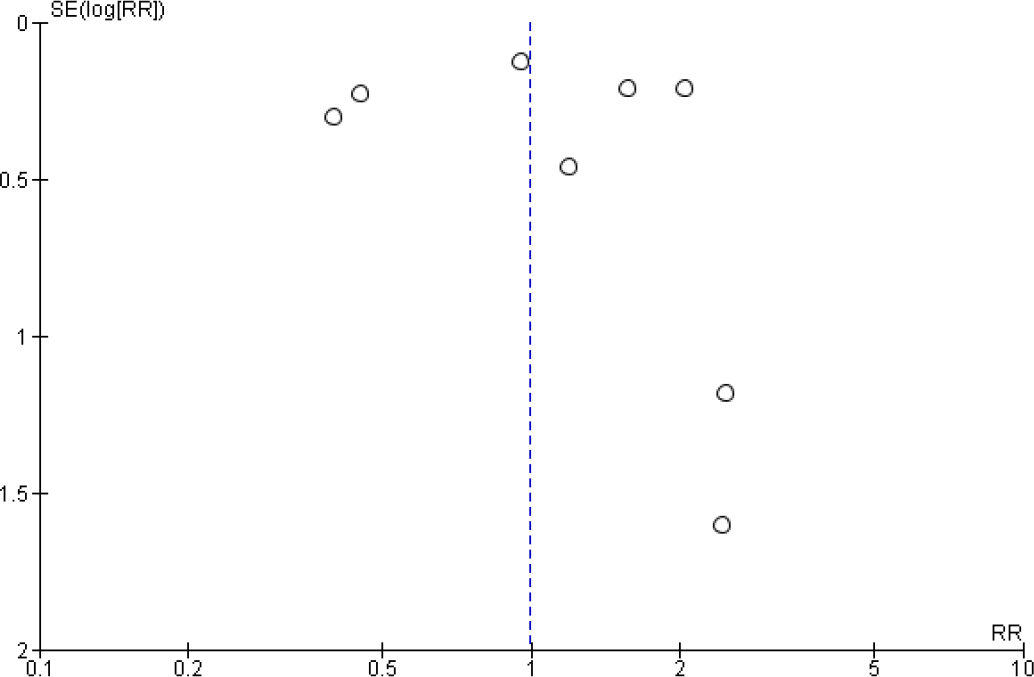
Funnel plot of included studies highlighted mortality of HCQ regimen.

### Hydroxychloroquine and Azithromycin Combination Versus Standard Care

#### Duration of Hospital Stay

The duration of hospital stay in case of treatment with (HCQ + AZM) combination versus standard care was reported in two studies. In the analysis, we found a significant difference between both groups where the (HCQ + AZM) combination group had the longer mean hospital stay among COVID 19 treated patients. The pooled Std. mean was 0.77, 95% CI, 0.46-1.08). The heterogeneity was statistically significant, P < 0.01, I^2^=92% (Figure 22).

**Figure 22.**
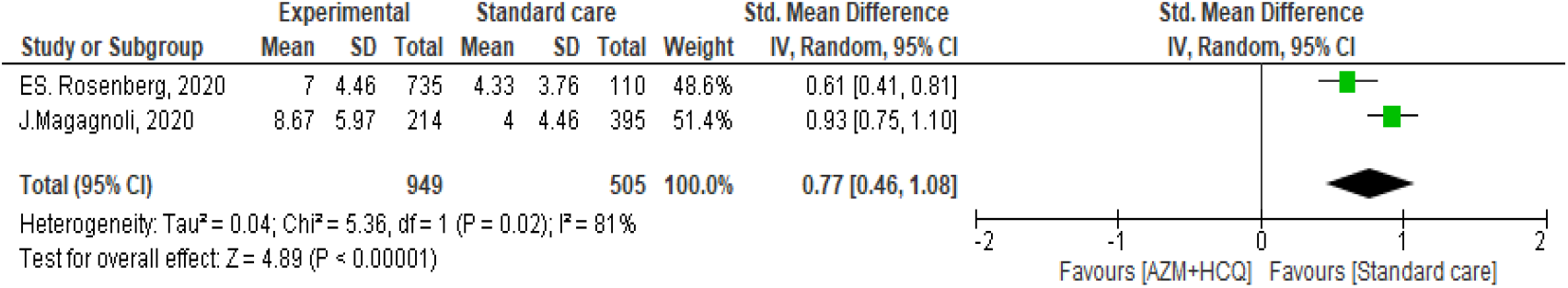
Duration of hospital stay of HCQ+AZM versus standard care.

#### Virological Cure Rate

Two studies reported the virological cure of HCQ+AZM combination versus standard care. The archived virological cure rate of HCQ and AZM combination (9/23) did not differ significantly from the standard care (4/31) (RR = 3.24, 95% CI, 0.71-14.74). The heterogeneity of the study was not significant P=0.12 I^2^= 58% (Figure 23).

**Figure 23.**
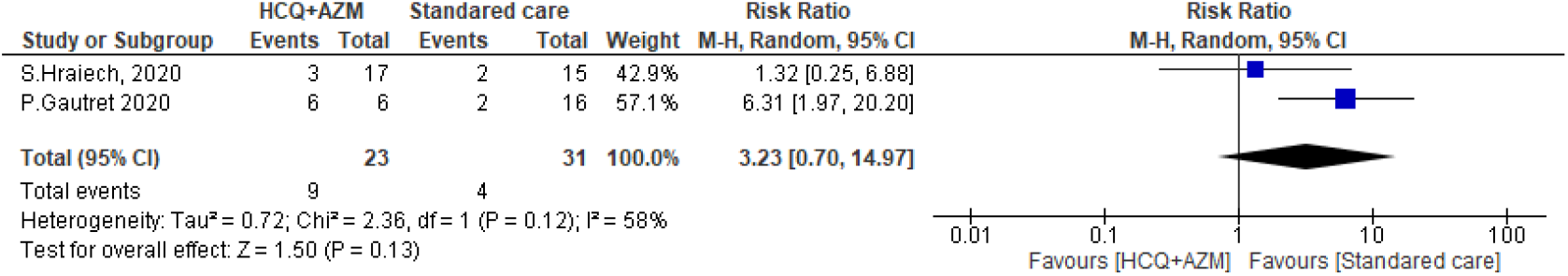
The virological cure rate of HCQ and azithromycin versus standard care.

#### Need for Mechanical Ventilation

Four studies evaluated the need for MV in both (HCQ+AZM) group (186/1627) and the standard care group (153/1389) and found no significant difference between both groups (RR = 1.27, 95% CI, 0.76-2.13). The heterogeneity of the studies was as follows I^2^= 88%, P < 0.01 (Figure 24). Leave one out sensitivity analysis was performed, ES. Rosenberg, 2020 contributed most to overall heterogeneity. By exclusion of this study, the heterogeneity between other studies was insignificant (P=0.82, I^2^=0%) (Figure 25).

**Figure 24.**
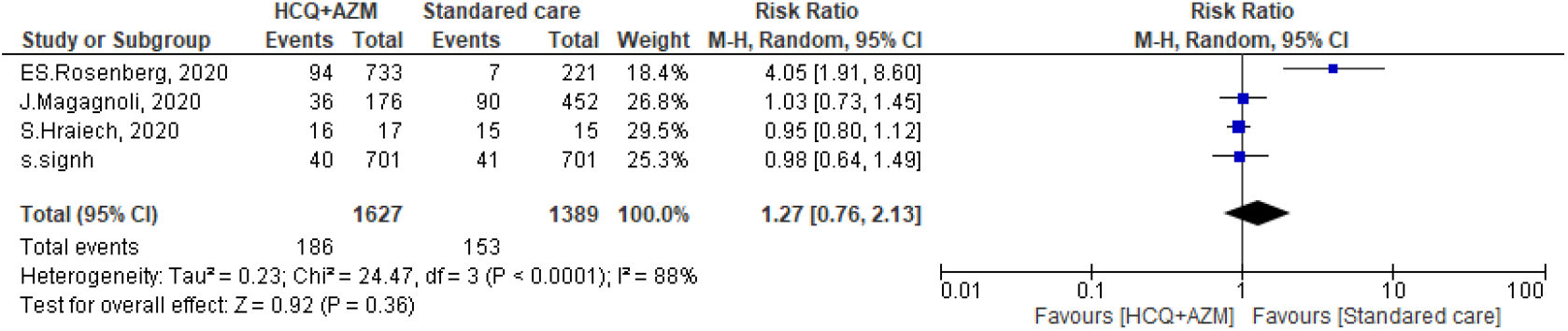
Need for mechanical ventilation of HCQ and AZM versus standard care.

**Figure 25.**
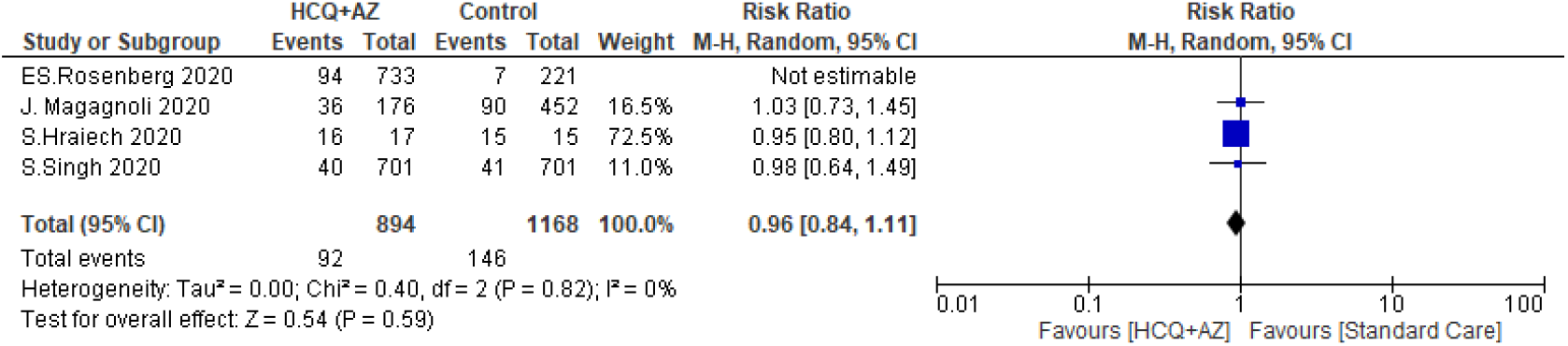
Leave one sensitivity analysis.

#### Mortality

Meanwhile comparing mortality in patients taking HCQ and AZM with those receiving standard care was addressed in 4 studies, two of them showed more mortality in the intervention group versus standard care group and the analysis there was no significant difference between the two groups with RR of 1.81, 95% CI, and 0.7-14.97 (Figure 26). Leave one out sensitivity test was performed. S. Singh, 2020 contributed most to the overall heterogeneity. By exclusion of this study, the heterogeneity between other studies was insignificant (P=0.71, I2=0%) (Figure 27).

**Figure 26.**
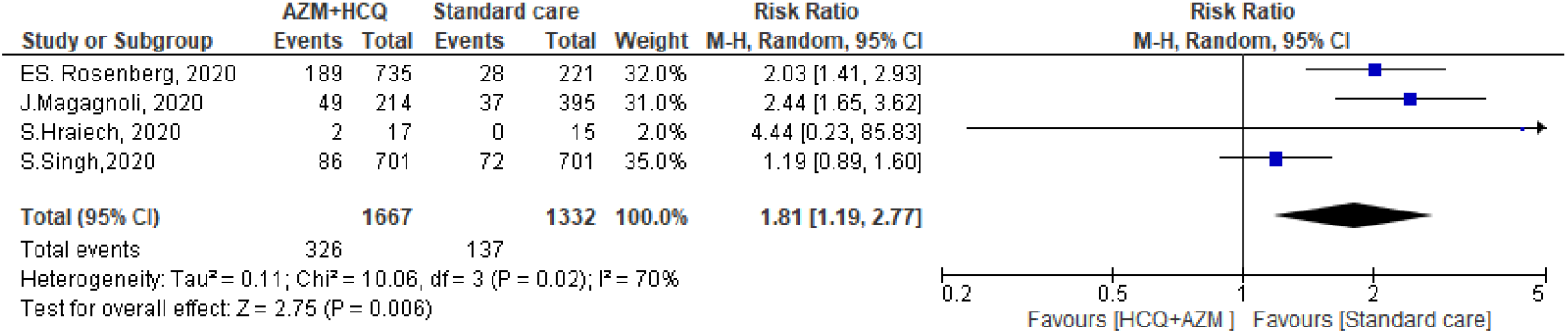
Mortality of Hydroxychloroquine and Azithromycin versus standard care.

**Figure 27.**
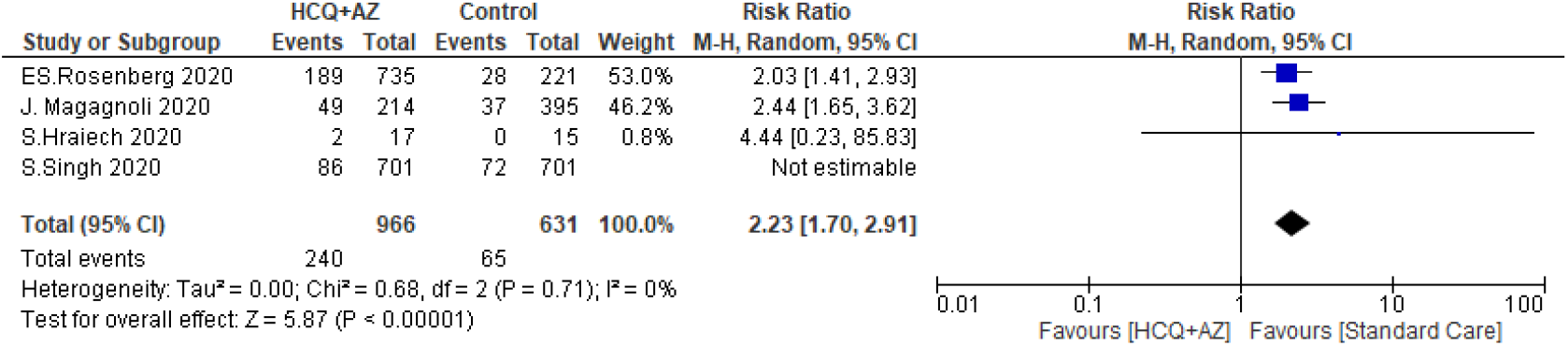
Leave one sensitivity analysis.

## DISCUSSION

Based on the finding of this meta-analysis, the treatment of COVID-19 patients with HCQ was associated with a longer duration of hospital stay, whether or not AZM was included in the treatment regimen. Regarding the difference in viral clearance between HCQ and standard care, time to negative conversion was not statistically different between the two groups (HCQ and standard care). Similarly, virological cure at either day 4, day 10, or day 14 was not different between both groups. It is worthy to mention that adding AZM to HCQ did not affect the cure rate versus standard care. Generally, exposure to HCQ alone or in combination with AZM wasn’t associated with the need for mechanical ventilation. Furthermore, neither clinical worsening nor radiological improvement of the studied patients was changed concurrently with exposure to HCQ. Side effects were more encountered if patients were exposed to HCQ, finally, the mortality of the HCQ group was not different from that of standard care. The country of residency was a significant predictor of mortality outcomes. The alarming finding was that the standard care had reported lower numbers of mortalities if they were compared to the AZM and HCQ combination group.

### Fever and Cough

Resolution of respiratory symptoms and fever is one of the symptoms-based indicators of disease recovery. In this study, we evaluated the recovery of these two symptoms after exposure to HCQ. ^(58)^ Huang et al, and Z. Chen et al demonstrated that patients treated with CQ recover from fever faster than those on standard care, however, J.Chen did not report any significant difference between both groups in the resolution of fever. Z. Chen et al reported a more significant resolution of cough among patients exposed to HCQ. Due to insufficient data, we did not conduct a meta-analysis.

### QT Prolongation

Abnormal myocardial repolarization results in QT interval prolongation. The normal QT interval is 470 ms in females, and 450 ms in males. ^(59)^ Among patients treated with HCQ, QT prolongation was identified in 23%-45.5%. ^(28, 37)^. About 12% of patients on AZM and HCQ stopped treatment due to significant QT prolongation. It is important to notice that there is no difference in the incidence of drug-induced QT prolongation by CQ or HCQ. A combination of HCQ and AZM increases the risk, however, Rosenberg et al, reported a lower occurrence of QT among patients received this combination versus standard care. ^(35)^

### Duration of Hospital Stay of HCQ ± AZM versus Standard Care

Adoption of either CQ/HCQ alone or in combination with AZM did not significantly shorten the duration of hospital stay. Patients on the standard care stay shorter in the hospital either if they compared to patient received CQ/HCQ (standard mean difference 0.54, 95% C10.20-0.94) or HCQ+AZM (standard mean difference 0.77, 95%CI 0.46-1.08) the reported heterogeneity of the CQ/HCQ study was 92% that dropped to 0% if the study of Huang was removed. While the heterogeneity of HCQ+AZM analysis was 81%

### Virological Cure Rate of CQ/HCQ ±AZM Versus Standard Care

In the current research, the achieved cure rate of HCQ (day 4, 10, and 14), and time to negative conversion among the HCQ group were not statistically different from the standard care. It is worthy to mention that, the term standard care was not firmly defined in each study, this may represent a source of prescription bias. Another important finding was that the pooled standard mean difference included the paper published by Huang et al, ^(3)^ this research resulted in significant heterogeneity in many outcomes especially time till virological cure. When we adopted the leave one sensitivity analysis the time to negative conversion became significantly shorter in the control arm.

Similarly, the virological cure rate of HCQ and AZM combination did not significantly differ from the cure rate of the standard care. This finding is similar to the result reported by Shamshirian et al,.^(60)^ In this study they included the study conducted by Maganogli et al, ^(41)^ in evaluating the effectiveness of HCQ+AZM combination. By reviewing this article, we found that the authors did not address the effectiveness of this combination versus standard care. ^(41)^.

### Radiological Improvement and Clinical Worsening

Treatment with HCQ did not provide any additional benefit in terms of radiological improvement or clinical worsening versus the standard care. We included three published articles in this analysis of the impact of HCQ on radiological improvement and five articles evaluating the effect of CQ/HCQ on clinical worsening. The heterogeneity of both analyses was 45% and 54% respectively.

### Need for Mechanical Ventilation of AZM+HCQ versus Standard Care

The need for mechanical ventilation was evaluated in our study in case of treatment with HCQ and (HCQ+AZM) combination compared to the standard treatment group. In our study, we included 5 studies in the comparison between HCQ and standard care, we analyzed the effect of (HCQ +AZM) combination in 4 studies and in both analyses no significant difference was found indicating that the use HCQ either alone or in combination with AZM for treatment of COVID 19 did not reduce the need for MV. Our results are in agreement with Shamshirian et al. ^(60)^ who published an MA of two studies addressing the need for MV among HCQ and standard care.

### Side Effects

In this meta-analysis, we included three published pieces of research that address the reported side effects of HCQ treatment. Patients on HCQ treatment had a higher risk of experiencing side effects, (pooled RR=3.14, 95% CI 1.58-6.24) with I^2^ of 0%. The reported side effects were diarrhea, headache, rash, elevated transaminases, fatigue, and anemia.

### Mortality HCQ and AZM

In the current research, the Mortality rate of HCQ alone did not significantly differ from the cure rate of standard care. This was similar to what is reported by Shamshirian et al, ^(60)^who almost included the same studies in his meta-analysis. The heterogeneity of this analysis was high so we conducted a sensitivity analysis to identify the source of heterogeneity. All studies contributed nearly equal to the reported heterogeneity, so we carried out meta-regression analysis. In Meta-regression analysis, the heterogeneity dropped to 22% and we identified that country was a strong predictor of mortality. One of the alarming findings was that the mortality of the HCQ and AZM was significantly higher than the standard care (pooled RR= 1.81, 95% CI 1.19-2.77). The heterogeneity of this analysis was (I^2^= 71%).

### Limitation

Our analysis must be interpreted in the context of the limitations of the available data; despite the huge number of published articles during the COVID-19 pandemic, many of these studies lack a good quality and may contain inconsistent results. In fact, there is an urgent need for high-quality randomized control trials that address the issue of HCQ treatment.

Consequently, we depended on our analysis of the few published or even cited preprints. Moreover, we included many observational studies due to the scarcity of randomized control trials. It is well established that observational studies cannot discover causality. This fact also contributed to the highly found heterogeneity of analysis especially for the study of Haung (2020). ^(3)^ After leaving one sensitivity analysis the heterogeneity drops to acceptable value in many outcomes. Another important source of bias in patient selection bias; as some studies did not classify patients according to their disease’s severity. This source of bias may significantly affect the course of illness. Differences in HCQ and AZM in dose, duration of treatment, and route of administration may also affect the consistency of our results.

## Conclusion

HCQ prolonged the duration of hospital stay and did not increase the overall virological cure or more specifically on days 4, 10, or 14. In addition, it did not affect duration till conversion to negative PCR, need for MV, radiological progression, clinical worsening of the disease, or death. Furthermore, treatment with AZM and HCQ did not affect the virological cure, the need for MV. However, it increased the duration of hospital stay and mortality. Future randomized clinical trials are needed to confirm these conclusions.

## Data Availability

The data is available and attached to this preprint

